# Individual-based modeling of COVID-19 transmission in college communities

**DOI:** 10.1101/2021.06.03.21258315

**Authors:** Qimin Huang, Martial Ndeffo-Mbah, Anirban Mondal, Sara Lee, David Gurarie

## Abstract

The ongoing COVID-19 pandemic has created major public health and socio-economic challenges across the United States. Among them are challenges to the educational system where college administrators are struggling with the questions of how to reopen in-person activities while prioritizing student safety. To help address this challenge, we developed a flexible computational framework to model the spread and control of COVID-19 on a residential college campus. The modeling framework accounts for heterogeneity in social interactions, activities, disease progression, and control interventions. The relative contribution of classroom, dorm, and social activities to disease transmission was explored. We observed that the dorm has the highest contribution to disease transmission followed by classroom and social activities. Without vaccination, frequent (weekly) random testing coupled with risk reduction measures (e.g. facial mask,) in classroom, dorm, and social activities is the most effective control strategy to mitigate the spread of COVID-19 on college campuses. Moreover, since random screening testing allows for the successful and early detection of both asymptomatic and symptomatic individuals, it successfully reduces the transmission rate such that the maximum quarantine capacity is far lower than expected to further reduce the economic burden caused from quarantine. With vaccination, herd immunity is estimated to be achievable by 50% to 80% immunity coverage. In the absence of herd immunity, simulations indicate that it is optimal to keep some level of transmission risk reduction measures in classroom, dorm, and social activities, while testing at a lower frequency. Though our quantitative results are likely provisional on our model assumptions, extensive sensitivity analysis confirms the robustness of their qualitative nature.

**Highlights:**

- Individual-based model for college communities with structured students’ interactions.
- We evaluated COVID-19 control measures needed for in-person college reopening.
- Without vaccination, high testing frequency is paramount for outbreak control.
- With high vaccination coverage, some NPIs are still needed for outbreak control.
- General access website tool was developed for the public to explore simulations.

Graphic Abstract

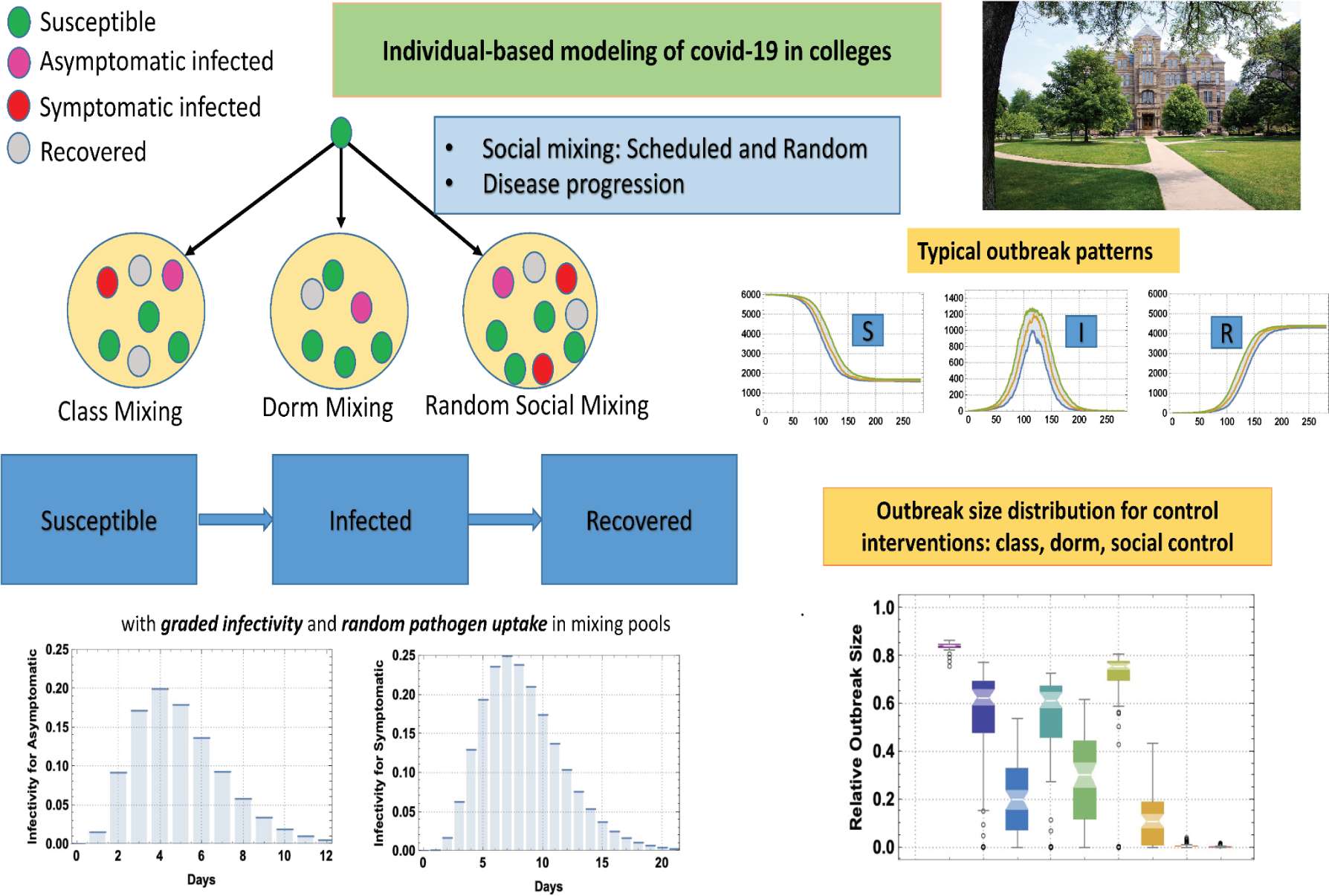

## Introduction

The ongoing COVID-19 pandemic has created major public health and socio-economic challenges across the United States. Among them are challenges to the educational system. After the 2020 Spring Break, many colleges and universities in the U.S. sent residential students home and transitioned partially or completely to remote learning as an attempt to reduce the risk of COVID-19 transmission on their campuses. Though such a policy may have contributed to reducing the spread of the disease on college campuses as well as their surrounding communities, it resulted in severe financial and potential academic losses for many colleges. In an attempt to mitigate these losses, college administrators are struggling with the questions of how to reopen in-person activities while prioritizing student safety. In August, 2020, over a third of US colleges and universities had fully in-person classes and around 20% chose to be hybrid (a mix of in-person and remote classes) [1]. Many of these reopenings were followed by campus outbreaks that led universities, including the University of North Carolina Chapel Hill (UNC) and the University of Notre Dame, and Indiana, to shut down or impose increased restrictions once again.

In the absence of vaccination and proven treatment, control strategies have focused on social/behavioral factors and preventive strategies such as facemask, reduced class size, reduced social gatherings/events size, and reduced residence hall (dormitory) occupancy to share the common utilities, bathrooms, and so on. Those strategies were combined with regular testing to rapidly detect and quarantine infected students. Previous modeling studies have investigated different aspects of COVID-19 spread on college campuses [2–7]. For example, Paltiel et al. [8] used a simple homogeneous mixing SEIR compartmental model to evaluate the effectiveness of screening strategies in a college with 5000 students; Weeden and Gornwell [9] designed student contact networks though transcript data and used these networks to studied the effectiveness of small course enrollments on reducing the risk of epidemic spread. Gressman and Peck [10] used a full-scale stochastic agent-based model (ABM) of a reasonably large research university to conclude that testing accuracy is a critical issue and holding large classes greatly increases the risk of a significant outbreak on campus. Goyal et al. [11] also developed an ABM that incorporates important features related to risk at the University of California San Diego to assess the potential impact of strategies to reduce outbreaks. They found that increased frequency of asymptomatic testing from monthly to twice weekly has minimal impact on average outbreak size, but substantially reduces the maximum outbreak size and cumulative number of cases. Inspired by their work, we developed an individual-based model to quantitatively assess the relative contribution from class, dorm, and social activities on the COVID-transmission and to quantitatively assess different control strategies such as screening and test with different frequency, class-size, dorm-size, and social-size control.

Even though vaccination of college students has started on most college campuses in the US, outbreaks could continue occurring due to incomplete population coverage and the appearance of new virus variants as the effect of vaccines on viral transmission and new virus variants are currently unknown [12, 13]. Given these uncertainties, university administrators are still determining to what degree they should continue adherence to non-pharmaceutical interventions.

### Challenges of Covid transmission and college specifics

Mathematical models are powerful tools to explore disease transmission, and the complex landscape of intervention strategies to quantify and assess different options. One of the key challenges in modeling the spread of Covid-like pathogens in local communities is an efficient account (parameterization) of human social interactions. Conventional approach (population-based) employs a simplified form of social interactions based on ‘mean contact rate/host’ [14, 15]. To address this limitation, individual-based modeling has been widely used in the study of pathogen spread and infection control, including Covid-19 [16–22]. Some have used individual-based models (IBM) where a community is viewed as a randomly colliding system of ‘host-particles’, whereby ‘infectives’ transmit the pathogen to ‘susceptibles’ via random collisions. The intensity (rate) of transmission is proportional to mean contact rate. On the opposite side are IBM based on social network structure where transmission pathways are confined to one’s ‘neighbors’ - adjacent nodes on the social network [23]. In real life human social contact mixing combines both random and scheduled (prescribed) contacts. Our proposed approach differs from previous works in two important aspects: 1) the basic transmission step in our model is ’many-to-many’ [24] vs. conventional ’one-to-one’; 2) we keep a detailed account of daily social interactions.

Here, we developed a flexible computational framework to model the spread of COVID-19 on a residential college campus. The modeling framework accounts for heterogeneity in social interactions, activities, disease progression, and control interventions. This allows us to generate and simulate more realistic synthetic college communities using a handful of basic inputs that combine scheduled and random contacts, and prescribed patterns of disease progression.

## Materials and Methods

Two basic components of college IBM include (i) college community and its social interaction patterns and (ii) infection transmission and disease progress. College community is made of individual student hosts (other population groups such as faculty and staff could be added). They are engaged in their daily activities (classes, dorm residence, social mixing). As a pathogen is introduced into such a community it could spread via social interactions.

I) **Student body and social mixing** is made of scheduled and random contacts (See Figure 1). Scheduled contacts include classes with assigned seating, and dorm residence. Random social mixing consists of daily aggregation in contact pools of different sizes and frequencies.
II) **Disease progression**. In our model, the infection stages are classified as susceptible (S), infected (I), and recovered (R) stages (See Figure 1). Once infected, each individual undergoes a prescribed infection progress pattern determined by stage duration and time-dependent infectivity (See Figure 2(A)). We partition the host population into two groups: asymptomatic and symptomatic. Each one undergoes SIR transitions of prescribed duration and pattern of the I-stage. In our setup variable (dynamic) infectivity serves as a proxy for viral load, a predictor for symptomatic expression, and diagnostic test sensitivity (See Figure 2(B)).

**Figure 1:**
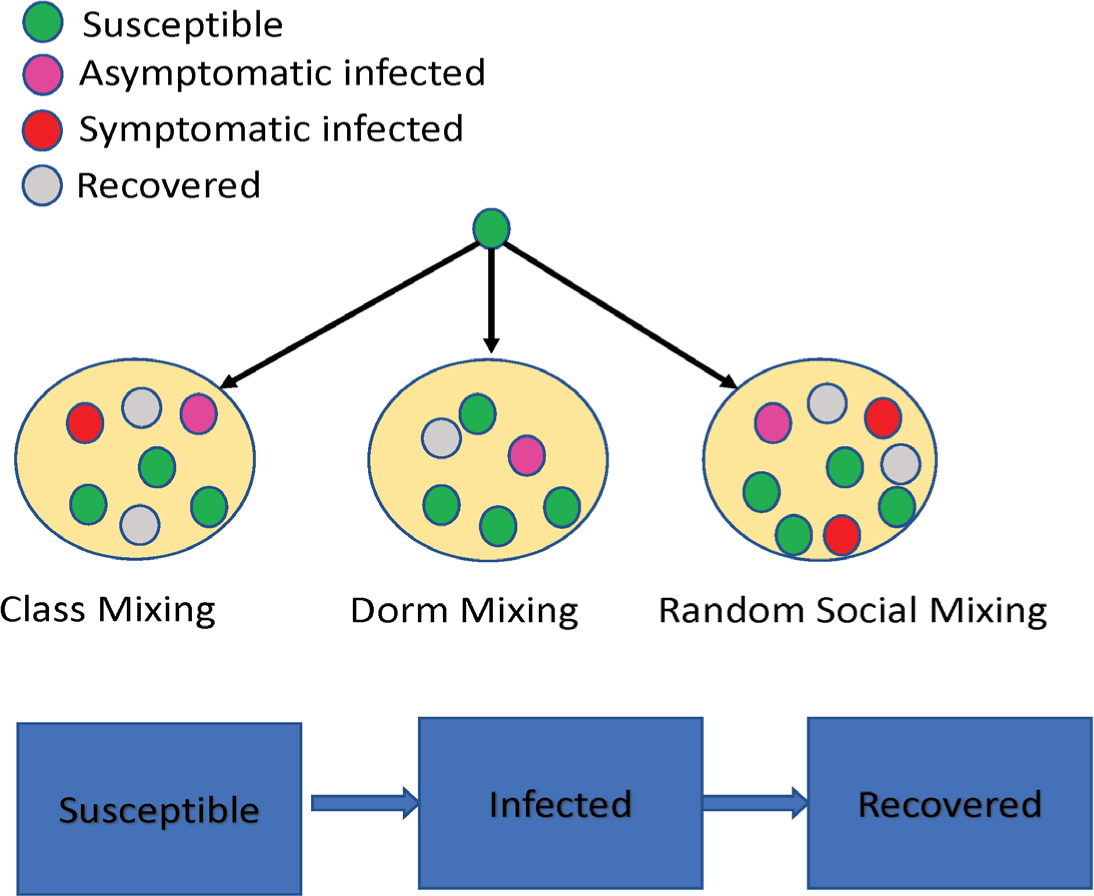
An abstract flowchart of social mixing and disease progress. Social mixing is implemented in our IBM via contact pools where students aggregate on a daily basis. Two types of contacts are considered: Scheduled (class and dorm mixing) and random (dining and/or other random social mixing). The infection stages are classified as susceptible (S), infected (I), and recovered (R) stages.

**Figure 2:**
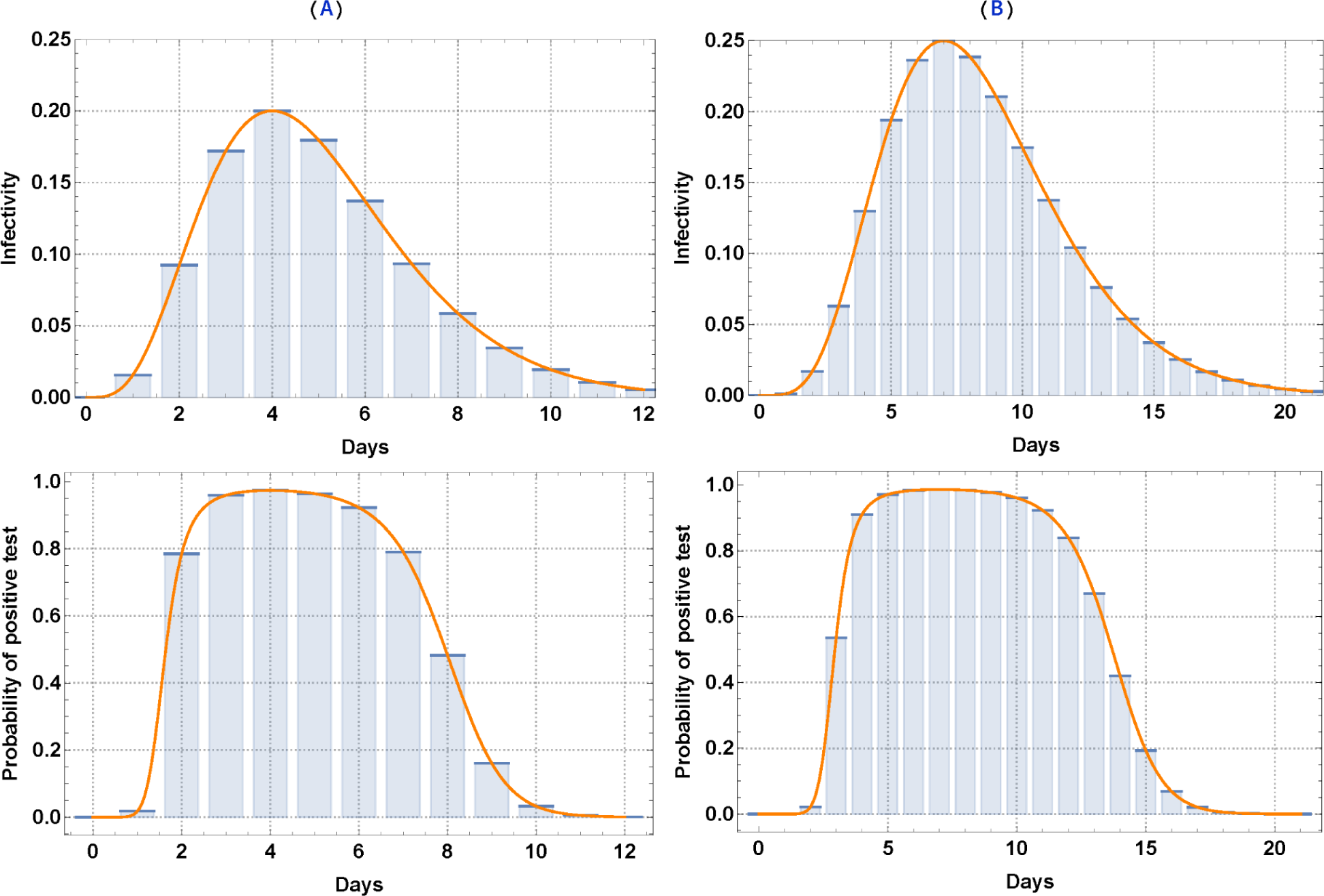
Upper panel is time-dependent infectivity levels for Asymptomatic and Symptomatic individuals, respectively. Lower panel is the time-dependent probability of a positive test for asymptomatic and symptomatic individuals, respectively.

### Social interactions

Social mixing is implemented in our IBM via contact pools where students aggregate on a daily basis. Two types of contacts are considered:

#### (i) Scheduled contacts

a. **Classes**. We generate a class schedule, including a seat assignment, for every student in our college. We assume that each student has three randomly selected non-overlapping classes. Classes are assigned to meet on a Monday, Wednesday, and Friday (MWF) schedule or on a Tuesday and Thursday (TuTh) schedule. All classes are assigned to a set of days and a time slot. Each class has a prescribed student seating in the classroom and weekly schedule. Students are assigned specific seats in each class period. Based on their seating pattern, an infected individual can spread the pathogen to his/her neighbors within a prescribed radius (Figure S1). Probability of transmission drops with longer distance.
b. **Housing/dorm.** A particular form of scheduled contact whereby students aggregate in shared units, halls, utilities etc. In our baseline setting, we randomly divide all students into non-overlapping groups with 7 occupancy per group.

#### (ii) Random social mixing

We simulate random social mixing using a specified pool size distribution. Specifically, students randomly aggregate on daily basis in contact pools of prescribed sizes *m*_1_; *m*_2_;…, and frequencies *f*_1_; *f*_2_;…. The latter accounts for an average number of mixing pools of a given type (per host per day), a proxy for their social-contact preferences. The real number of mixing pools of size *m_i_* is equal to *Nf_i_* (*N* -total student body size). As each m-pool creates *m*(*m* −1) daily contacts, the mean contact rate /host is equal to *w* = ∑ _*m*≥2_ *f_m_m*(*m* −1) (see Table S1). The mean contact rate plays an important part in the standard SIR modeling, but in our setting the key inputs are numbers {*m*_1_; *m*_2_;…};{ *f*_1_; *f*_2_;…}. More refined contact pools can be generated from the social network makeup of the host community.

#### (iii) Transmission environment

Student mixing pools (scheduled and random) aggregate on a daily basis, and provide the key transmission mechanism from infected to susceptibles. Furthermore, a host could partake in several daily activities, and thereby accumulate his/her exposure, or contribution to disease spread. We assume each type of activity (class, dorm, and social) takes place in a suitable environment, and each environment has an has an associated mean risk factor (*a_c_*, *a_d_*, *a_s_*). These factors (0 < *a_i_* < 1) mitigate the probability of transmission (exposure risk) in a given environment, They vary from 0 (full protection) to 1 (full exposure). In this form they could also be the target of control strategies. More generally, risk factors could be made site-specific (classroom and/or other place of activity). Combined with individual infectivity, environmental risk factors determine the probability of I ->S transmission as elaborated in the next subsection.

Note that the risk factors (*a_c_*, *a_d_*, *a_s_*) associated with three transmission environments (class, dorm, and social) are highly uncertain and could vary in different colleges. Hence we explore a range of choices of environmental risk factors: three choices (low, moderate, high) for three environmental risk factors; Besides, we also consider two choices of random social mixing cluster sizes (mean contact rate (MCR) =3, MCR=6). Other parameters of disease progression are reasonably chosen based on literature (See Table S3).

**Table 1:**
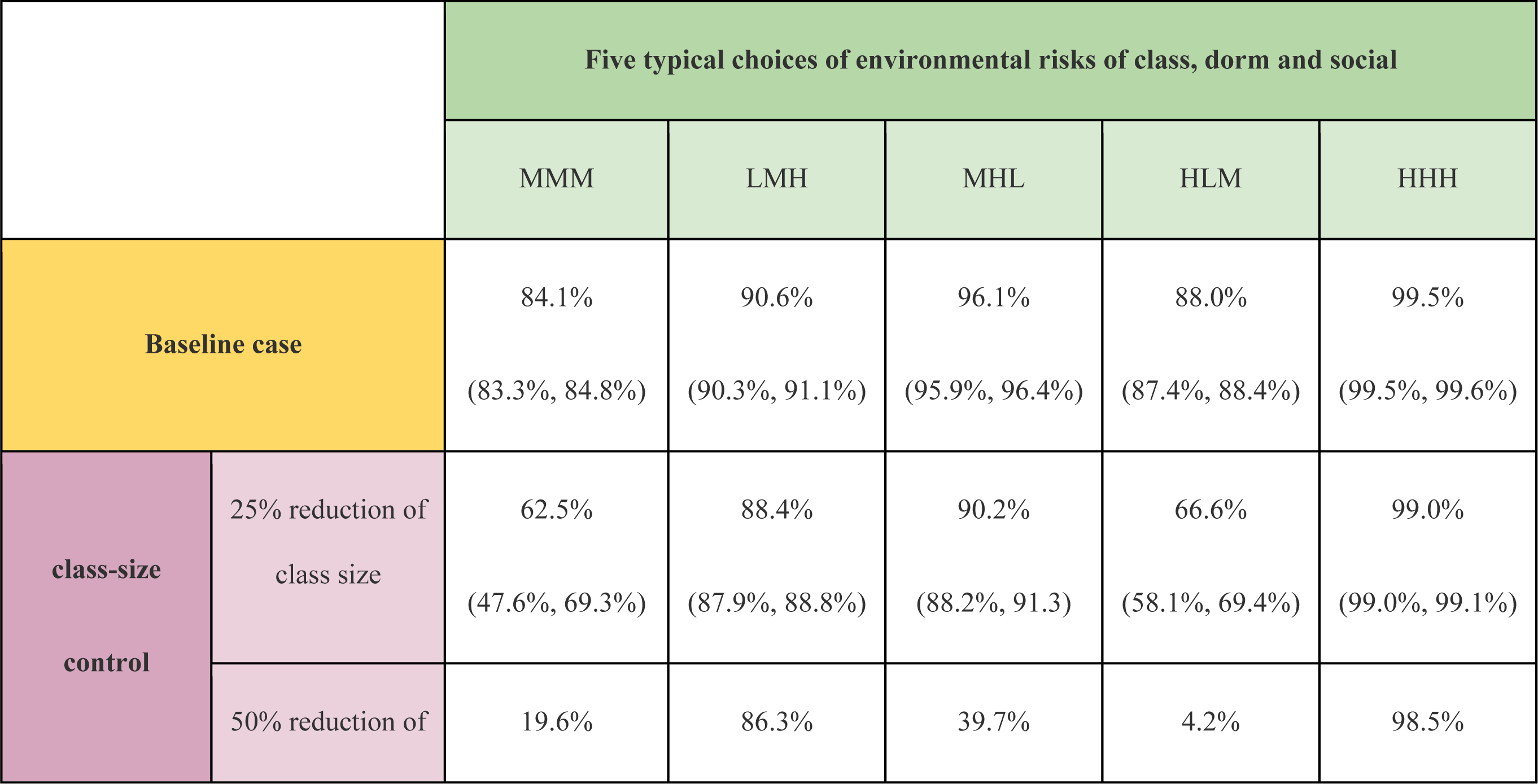
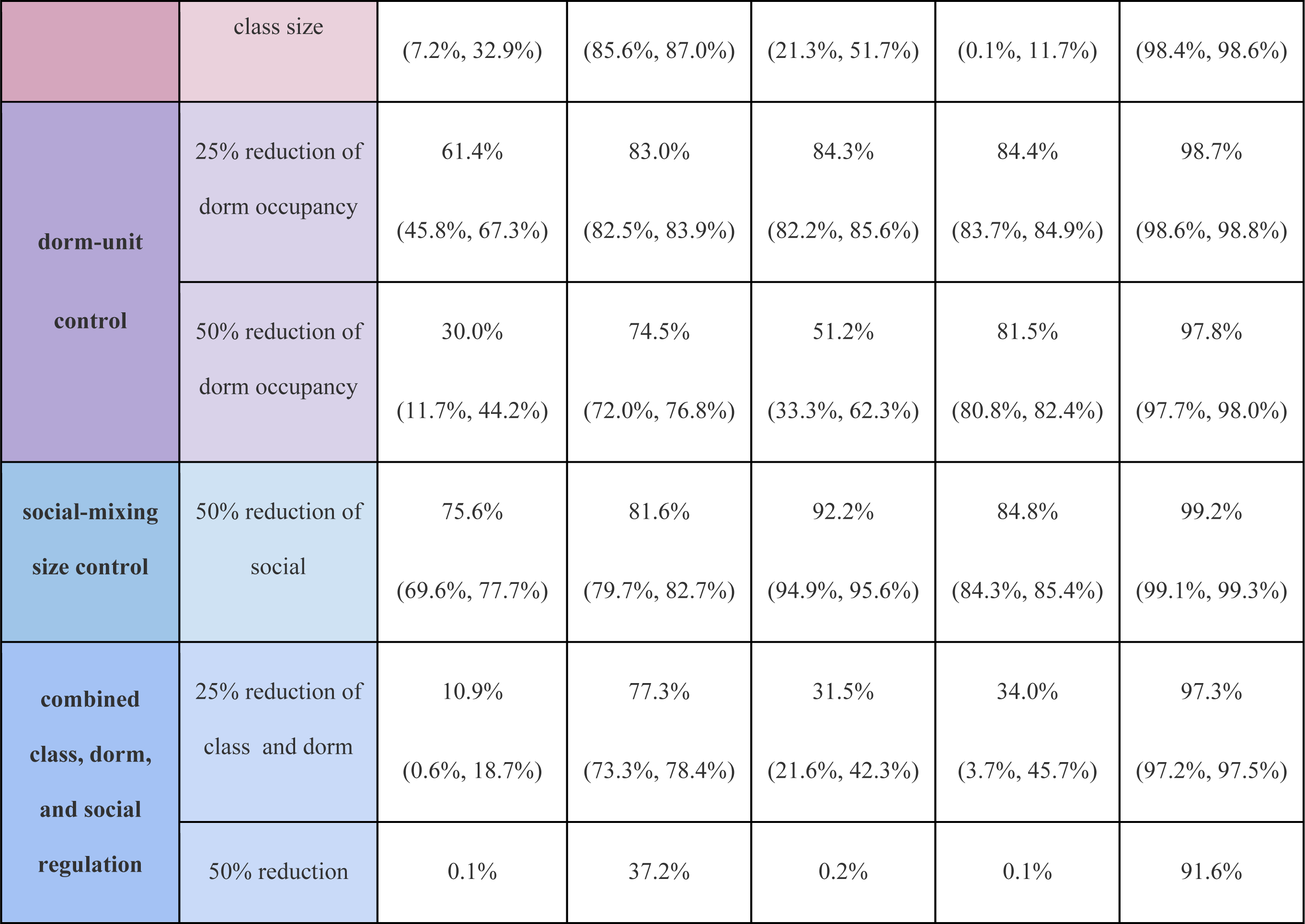
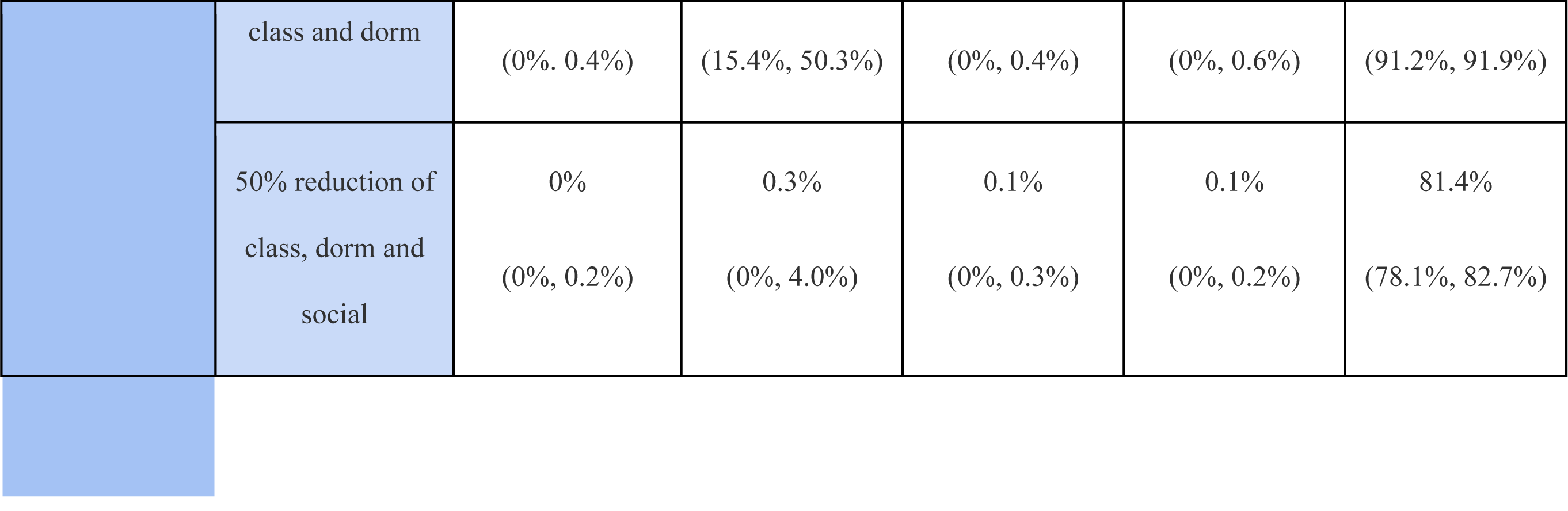
The effect of class, dorm, and social size on relative outbreak size with a 6000 student body during a 16-week semester. We selected five typical cases as our baseline cases: MMM (the first letter represents the risk level for class, followed by dorm and social), LMH, MHL, HLM, and HHH. For example, LMH means low risk factor for class, moderate risk factor for dorm, and high risk factor for social activity. We explored the 8 different choices of class, dorm, and social size regulation. The results shown are predicted median (25% quantile and 75% quantile)

### Infection and disease progress

Different hosts can undergo different disease processes in the I-stage. Here for simplicity we partition the host population into two groups: asymptomatic or symptomatic. The key difference between two is the stage duration and the infectivity function profile. In our setup asymptomatic hosts have shorter duration and lower infectivity [25, 26]. Asymptomatic hosts do not exhibit symptoms over the duration of I-stage, but are still capable of transmitting the pathogen. In contrast, Symptomatic hosts would exhibit disease symptoms after their incubation period.

The key quantifier of disease progress in the I-stage, is host infectivity function or post infection *b*(*d*) day *d*. We view it as a proxy for variable viral load in the course of infection. It follows a prescribed dynamic pattern (shown in Figure 2(A)); specifically we use function *B*(*t*) = *t ^k^e*^−*kt*^ of steepness *k*, and define *b*(*d*) = *b_M_ B*(*d/L*) where *b_M_* is peak infectivity and *L* is the mean duration. Each I-host would progress according to his/her *b*(*d*), and such patterns could be made individual. But in the current version we partitioned host population into two groups: asymptomatic and symptomatic with different parameters choices for (*b_M_*; *k*; *L*) (see Table S3).

The infectivity function *b*(*d*) determines the probability of transmission per contact from an infected I-host to a fully susceptible S-host. Hence, we employ it to estimate transmission rates within contact pools. We also use it (timing *d*) as a predictor for symptomatic isolation, and values of *b*(*d*) for predicting positive test diagnostics in screening (See Figure 2(B)).

Host infection (*S* → *I*) is modeled as a Bernoulli random process with the success probability determined by cumulative daily contact pools of a given susceptible -host. In each contact pool, scheduled or random (class, dorm, social mixing), we estimate the survival probability (staying uninfected) depending on the pool’s makeup, and the associated-environmental risk factor.

Specifically, a contact pool with *m* infectives with infectivity levels {*b*_1_;…;*b_m_*}, and environmental risk factor *a* gives a survival probability of an S-host, 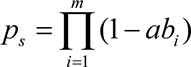. Note that the classroom setting is somewhat different from other contact pools, as the probability of survival depends not on the entire body of infected classmates, but only on the ‘nearest infected neighbors’ determined by classroom seating (see SI for details). Several contact pools for a given S-host, could accumulate over a daily period; they determine the final probability of infection, *p_I_* = 1− *p_sc_* · *p_sd_* · *p_ss_*, where *p_sc_*, *p_sd_*, *p_ss_* are survival probabilities in class, dorm and random social mixing.

### Control interventions

Our IBM setup allows us to investigate a wide range of interventions, including (i) symptomatic isolation, (ii) scheduled and random test screening at prescribed weekly schedule; (iii) regulation of class size, dorm occupancy, or social mixing makeup; (iv) vaccination. Furthermore, each transmission environment (classroom, dorm, social) has an associated environmental risk factor which affects the probability of transmission in the setting. These risk factors account for physical conditions (e.g. seat density in classrooms, specifics of dorm units), and behavioral practices (e.g. adherence to face mask or other protective means during contacts). Such environmental inputs are highly uncertain and we explore a range of choices to assess their impact on disease outbreaks and the effectiveness of control strategies.

Different control strategies can be compared in terms of their efficacy in mitigating an outbreak. For quantitative assessment of control interventions and their impact, we use two measurements: (i) outbreak size (the cumulative infection by the end of a given time period (e.g. 16-weeks semester) relative to total population); (ii) daily maximal quarantine number estimated from the quarantine pool over the outbreak period. The latter could provide some guidelines to the University administration on how many quarantine rooms to prepare in advance, and could also give a simple economic measure of outbreak impact and putative intervention. Those are estimated and compared with baseline cases to assess the efficacy of control.

### Numeric procedures, key inputs, and website tool developed

All codes of the college IBM and the tools of analysis were implemented in the Wolfram Mathematica platform. Supplement gives a specific parameter used in model simulations, and an outline for generating a synthetic college community from typical class size enrollment, living accommodations, and social activities. Overall, it consists of four key parts: (i) class, dorm, social interaction, and environment risk factor; (ii) disease progression parameter; (iii) control parameter, (iv) student body and initial conditions. A friendly-used website tool was also developed for the public to explore the simulation https://www.wolframcloud.com/obj/ait14/ibmcollegecovidmodel.

## Results

### Outbreak in naive host community

We generate a synthetic medium-sized (6000) and a small-size (3000) college community with all students residing in dorms: no intervention (only depends on disease, social mixing, and environment), no pre-existing immunity, and no external source of infection. All results with the 6000 student body are represented here, and with the 3000 student body are put in (See Figures S3-S8 in SI). A naive host population is initialized with a few infected individuals.

### Typical outbreak patterns and parameter sensitivity: baseline cases

A typical outbreak pattern and duration depends on transmission intensity. Under high transmission intensity, outbreaks are localized within a limited range (typically less than a 16-week academic semester). However, if transmission intensity is low, they could last much longer and won’t be terminated by the end of semester (See Figure 3).

**Figure 3:**
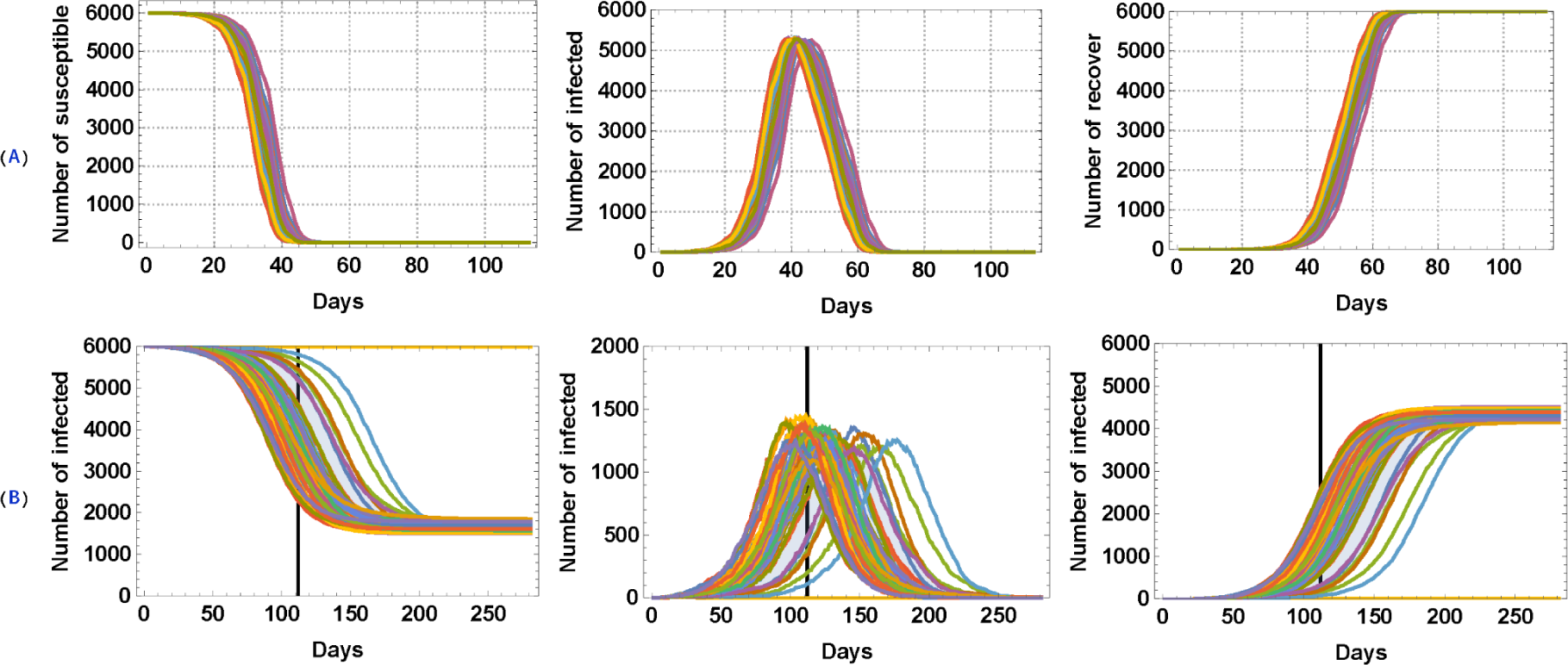
Typical outbreak histories of susceptible, infected, and recovered for a high transmission scenario (Panel A) and for a low transmission scenario (Panel B), respectively. The black vertical line in Panel B means the end of a 16-week academic semester.

All our college simulations were run over a 16-week academic semester. Under high transmission intensity (e.g. high environmental risk), almost all students got infected by the end of a 16-week semester. Therefore, the outbreak size distribution is localized (shown in Figure 3(A) and Figure 4). However, low transmission intensity creates a highly dispersed statistical outcome by the end of the 16-week semester, as not all random histories are terminated (See Figure 3(B) and Figure 4). The dispersal also grows larger for smaller student bodies (see details **Figure S3** in SI, for a 3000 student body).

**Figure 4:**
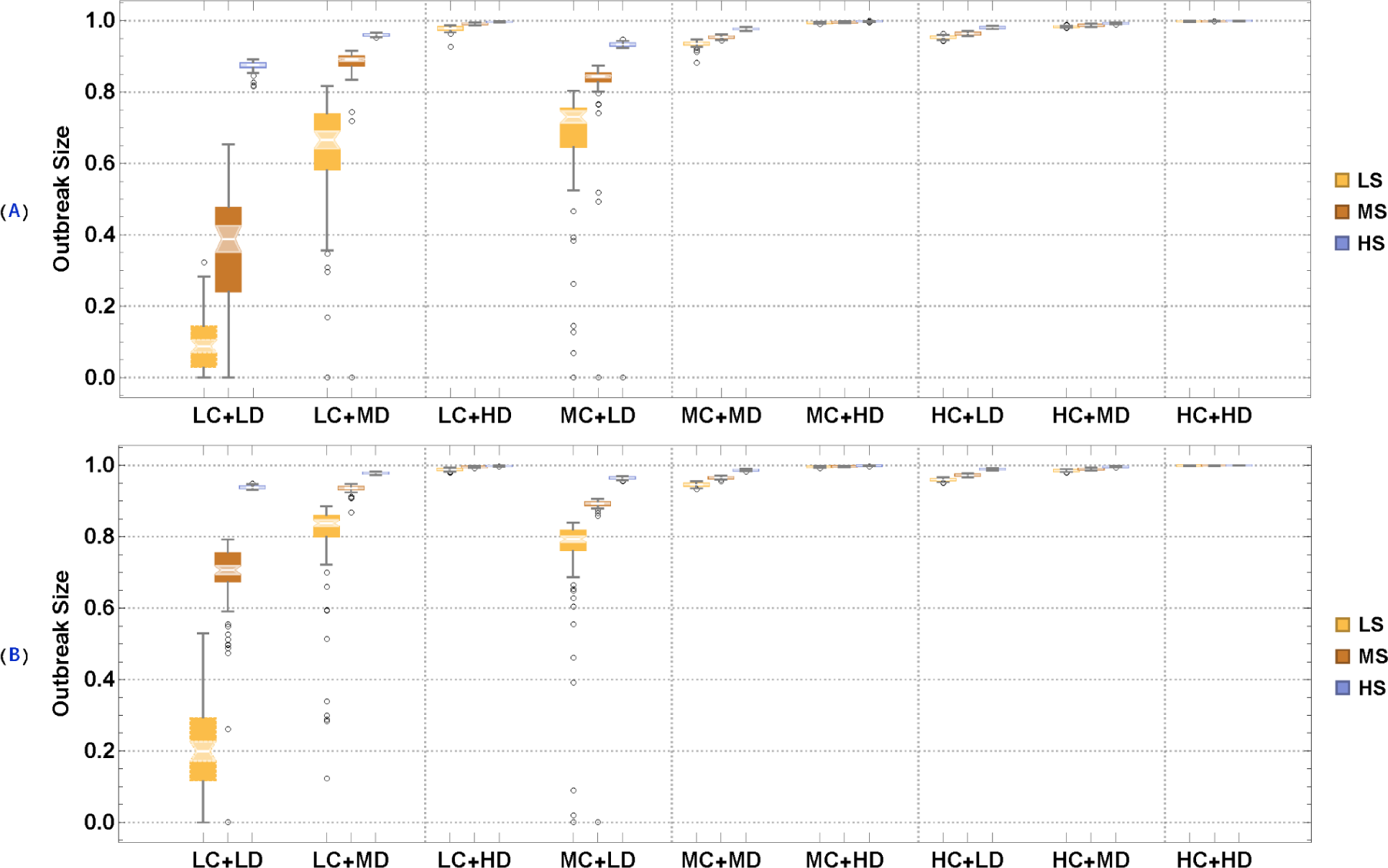
We consider three different levels of environmental risk (low, medium, and high) for three environments (class, dorm, and social mixing) and two choices of social mixing sizes (MCR=3 and MCR=6) to run our college IBM for a 16-week semester. LC, MC, and HC represent low, medium, and high risk factors for class; LD, MD, and HD represent small, medium, and high risk factors for dorms; LS, MS, and HS represent small, medium, and high risk factors for social activity. The output is split into triples, so we have 27 combinations of parameter choices for each panel. 100 realizations were run for each case and the light shaded region indicated the quantiles of the distribution of the median. Panel (A): random social mixing: mean contact rate (MCR)=3. Panel (B): MCR=6 case.

To compare Panel (A) with Panel (B) in Figure 4, we can see that random social mixing size also plays an important role in controlling the spread of COVID-19 in college students. From now on, the following experiments are implemented in the setting of the mean contact rate (MCR) of random social mixing =6.

To further explore and assess the relative contribution of three environments (class, dorm, and social) on outbreak size, we fixed other parameter values to see what would happen if one environment is ignored, as shown in Figure 5. We observed the relative contribution of the dorm was most significant followed by class and social mixing. Such conclusions could be provisional on our assumptions about dorm, class, social make-up.

**Figure 5:**
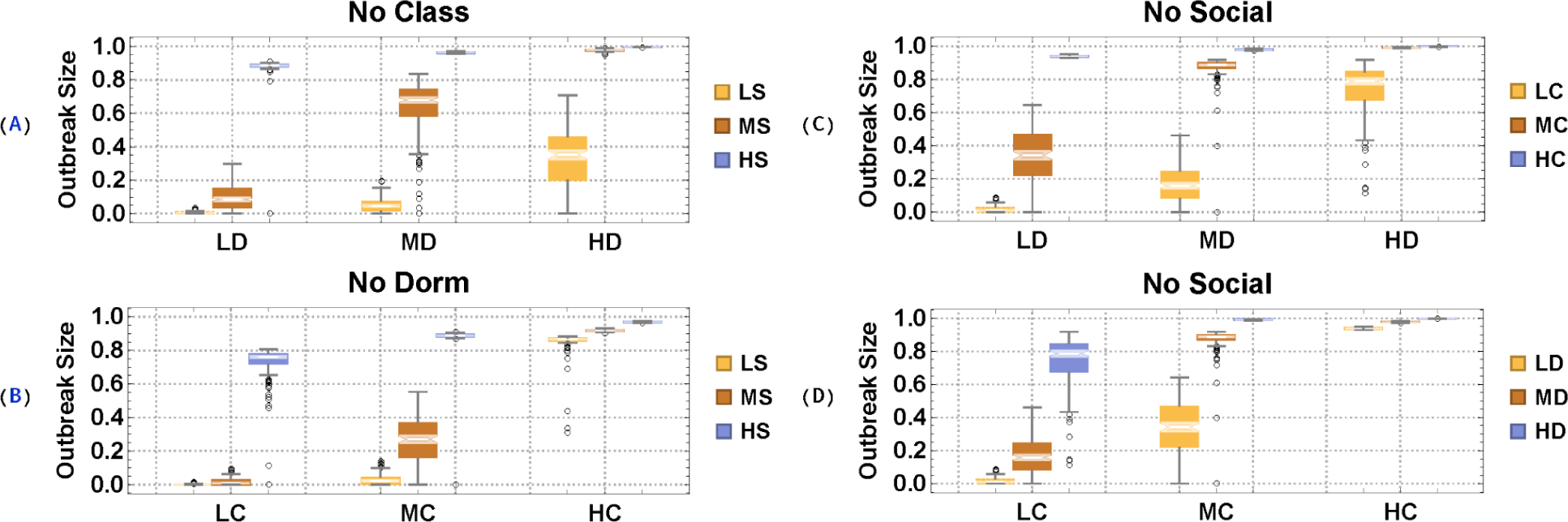
Predicted outbreak size distribution obtained by removing specific transmission environments: panel (A) no classroom transmission; Panel (B) no dorm transmission. Panel (C) and (D) no random social mixing. Mean contact rate for random social mixing in all cases is MCR=6.

### Analysis of intervention

#### Baseline case with symptomatic quarantine

We start with a simple scenario (symptomatic quarantine only) shown in Figure 6. We assume that an infected individual would start showing symptoms at around 7 days, based on our time-dependent infectivity level for symptomatic pathways shown in Figure 2. 70% random selection of those individuals showing symptoms are put into quarantine, where they do not mix and transmit the pathogens. Compared with Figure 4 where no control intervention applied, the symptomatic quarantine only scheme (Figure 6) may work fine for the case that we have really good control of risks from all three environments (class, dorm, and social), but definitely not enough for those cases with moderate and high risk factors. In the latter, the scheme has minimal impact on the outbreak size, but raises a significant economic burden to do quarantines.

**Figure 6:**
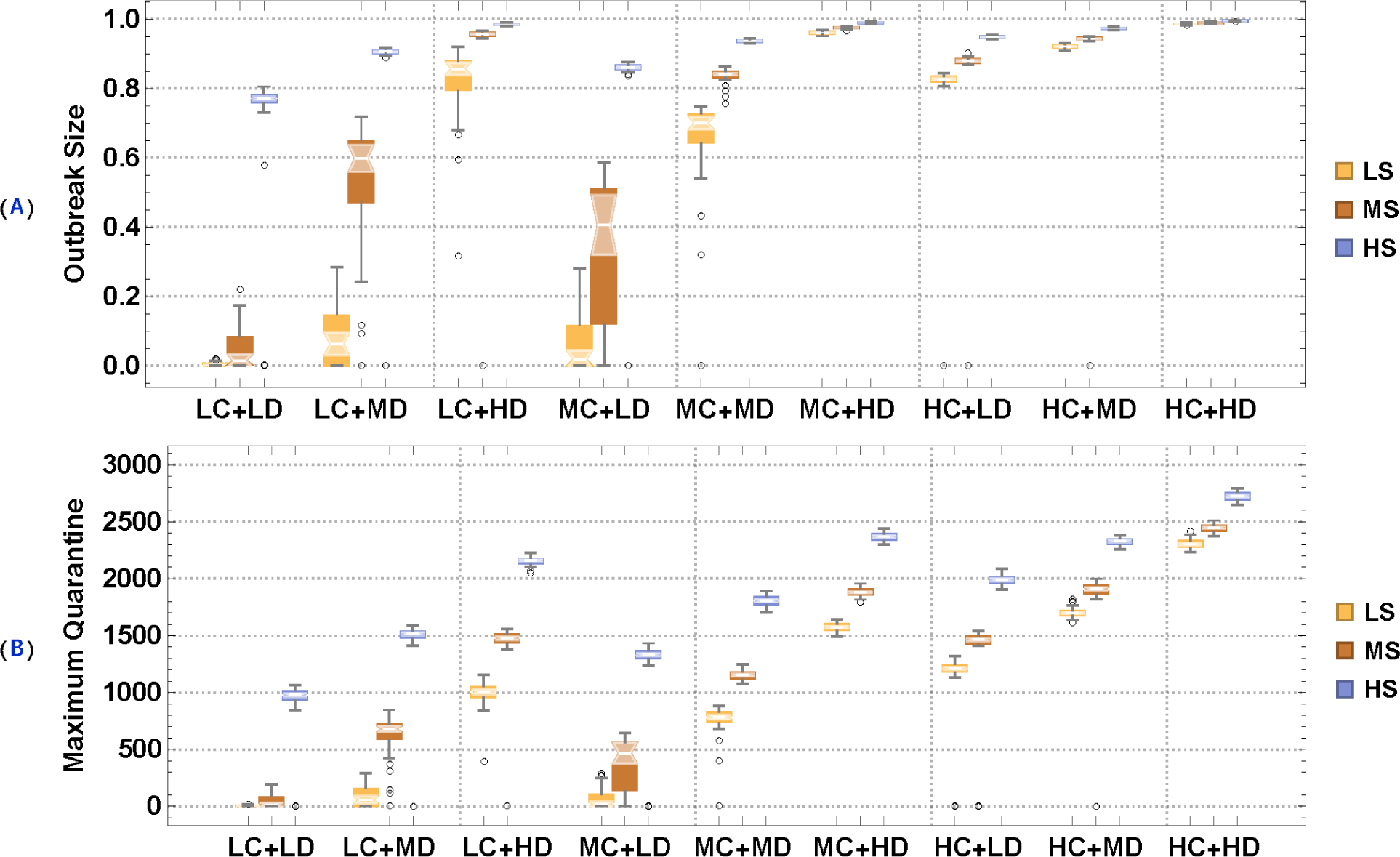
The effect of symptomatic quarantine on relative outbreak size and maximum quarantine capacity (for college size 6000). We assume an infected individual from a symptomatic pool would exhibit symptoms after day 7, based on our time-dependent infectivity pattern (Figure 2). A random fraction of those (70%) are quarantined and don’t contribute to transmission. Once recovered they are released into the normal (class-dorm-social) setting.

The clue to low efficacy of symptomatic screening lies in (i) transmission rate is still high, though symptomatic quarantine tries to lower it; (ii) the role of asymptomatic individuals in transmission and the delay of detection. To test (i)-(ii) in the following subsection, we first selected five typical cases from Figure 6 (70% symptomatic quarantine only) as our baseline cases: MMM (the first letter represents the risk level for class, followed by dorm and social), LMH, MHL, HLM, and HHH. For example, LMH means low risk factor for class, moderate risk factor for dorm, and high risk factor for social activity.

#### Class, Dorm, and Social size regulation

We then consider the following means of social control: reduced class size, dorm occupancy, random mixing pools, coupled with 70% symptomatic quarantine: (i) class size reduction at 25% and 50% is achieved by randomly selecting a fraction of students to attend in-person from the original class enrollment. While removed from their classes the remaining fraction could still contribute to transmission through other activities; (ii) dorm occupancy per unit is similarly reduced by 25%, 50%; (iii) random social mixing is reduced by 50%, via regulating pool sizes and their frequency; (iv) combined interventions: (a) 25% class-size reduction + 25% dorm-occupancy per unit reduction, (b) 50% class-size reduction + 50% dorm-occupancy per unit reduction, (c) 50% class-size reduction + 50% dorm-occupancy per unit reduction + 50% mean contact rate of random social mixing reduction.

Figures 7-8 and Table 1 show the effect of above interventions on relative outbreak size and maximum quarantine capacity, respectively. The last scheme, 50% class-size reduction + 50% dorm-occupancy-per-unit reduction + 50% mean contact rate of random social mixing reduction, could prevent an outbreak with some control on environmental risks, but still would lead to failure if there is no control on environmental risks. Overall, the effect of class, dorm, and social mixing size regulation is highly dependent on the risk control (e.g., wearing facial masks in class, dorm, and other social activities) in those three environments. Moreover, the reduced transmission would lead to a lower number of quarantine rooms, which reduces the economic burden of the University.

**Figure 7:**
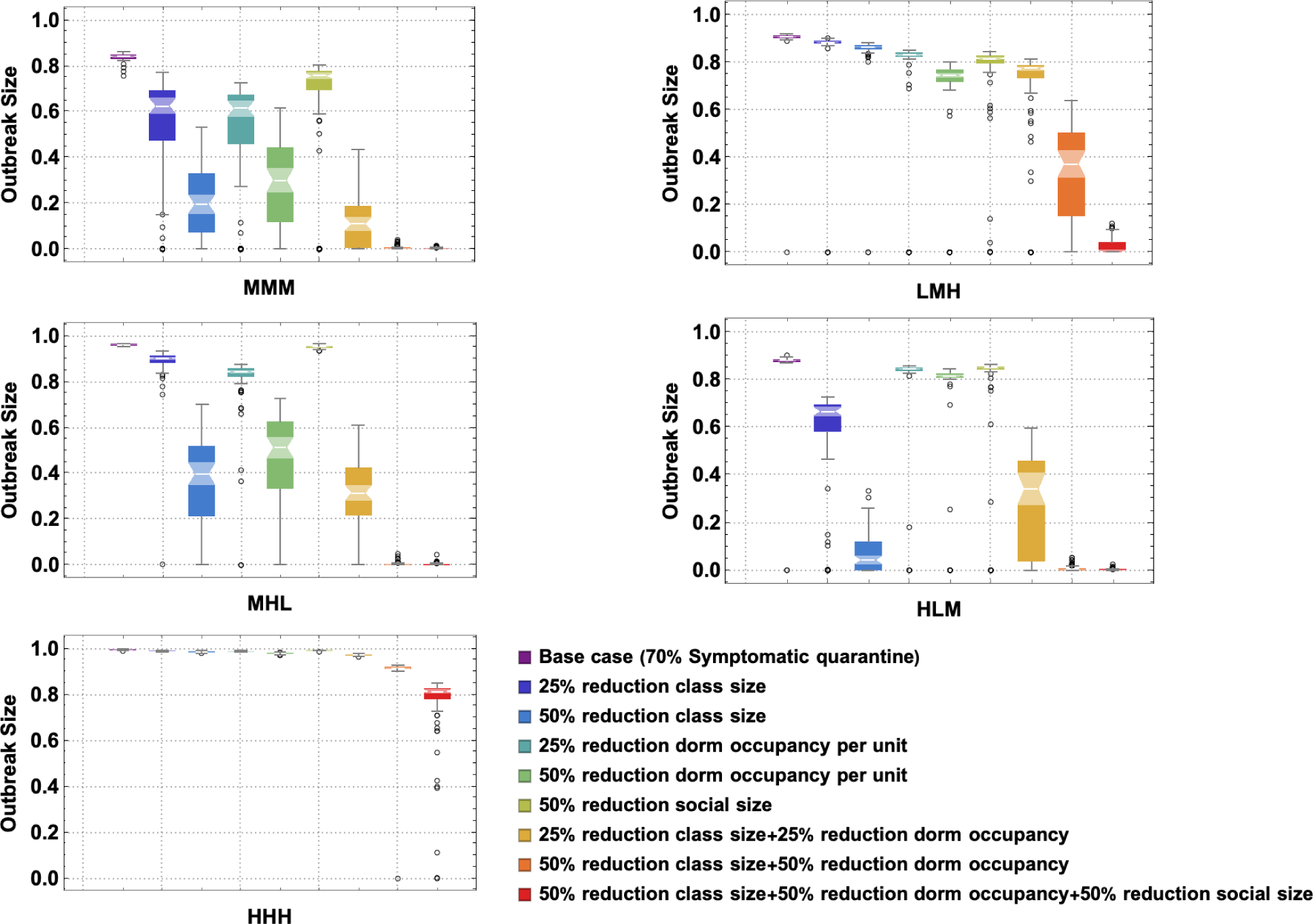
Combined control interventions of class-size, dorm-occupancy, and social mixing pools on outbreak size. We selected five typical reference cases: MMM (the first letter represents the risk level for class, followed by dorm and social), LMH, MHL, HLM, and HHH. For example, LMH means low risk factor for class, moderate risk factor for dorm, and high risk factor for social activity. We explored the 8 different choices of class, dorm, and social size regulation.

**Figure 8:**
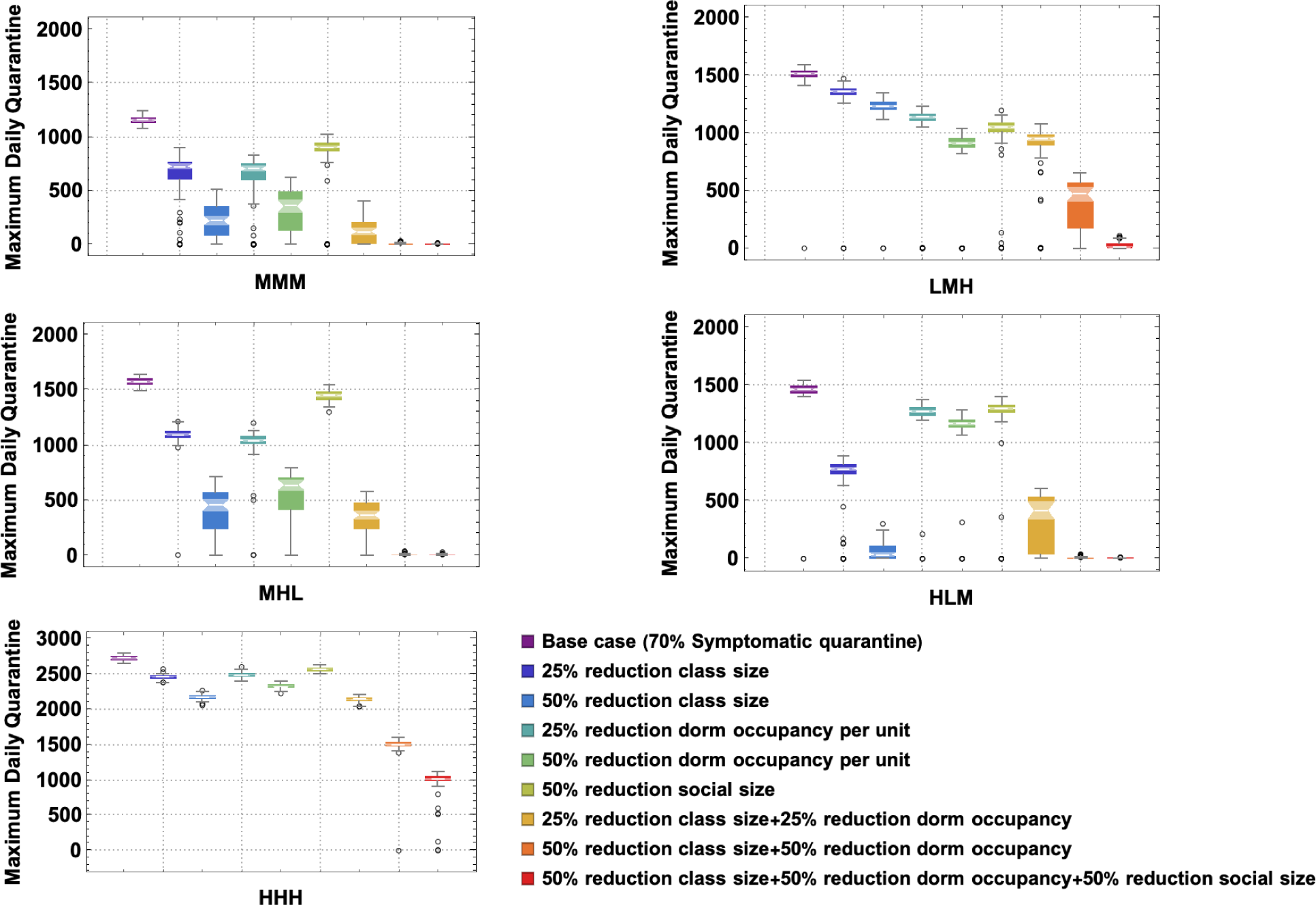
The effect of different combinations of class, dorm, and social control on maximum quarantine. We selected five typical cases as our baseline cases: MMM, LMH, MHL, HLM, and HHH, using the same setting as in Figure 7. We explored the 8 different choices of class, dorm, and social size regulation.

#### Screening and Tests

Next, we explore different Screening and Test policies coupled with 70% symptomatic quarantine as follows: (i) we assume that the University is capable of doing about 7% of population screening capacity and tests per day from randomly selected pools (i.e., about 100% of the population every two week). The sensitivity of the test is assumed to be time-dependent for both asymptomatic and symptomatic infected individuals (Figure 2); (ii) we assume that the University is capable of doing 14% of population screening and tests per day from randomly selected pools (i.e., about 100% of the population per week).

Note that the above two Screening and Test policies are coupled with daily random 70% symptomatic quarantine (i.e., an infected individual is assumed to start showing symptoms at around 7 days of infection, then 70% random selection of those individuals showing symptoms are put into quarantine).

Figure 9 and Table 2 show that the high frequency of screening and tests is an effective control strategy to mitigate and even prevent the outbreak of COVID-19 in colleges coupled with some control of the risk factors in class, dorm, and social environments. Moreover, since random screening testing allows for the successful and early detection of both asymptomatic and symptomatic individuals, it successfully reduces the transmission rate such that the maximum quarantine capacity is far lower than expected to further reduce the economic burden caused from quarantine.

**Figure 9:**
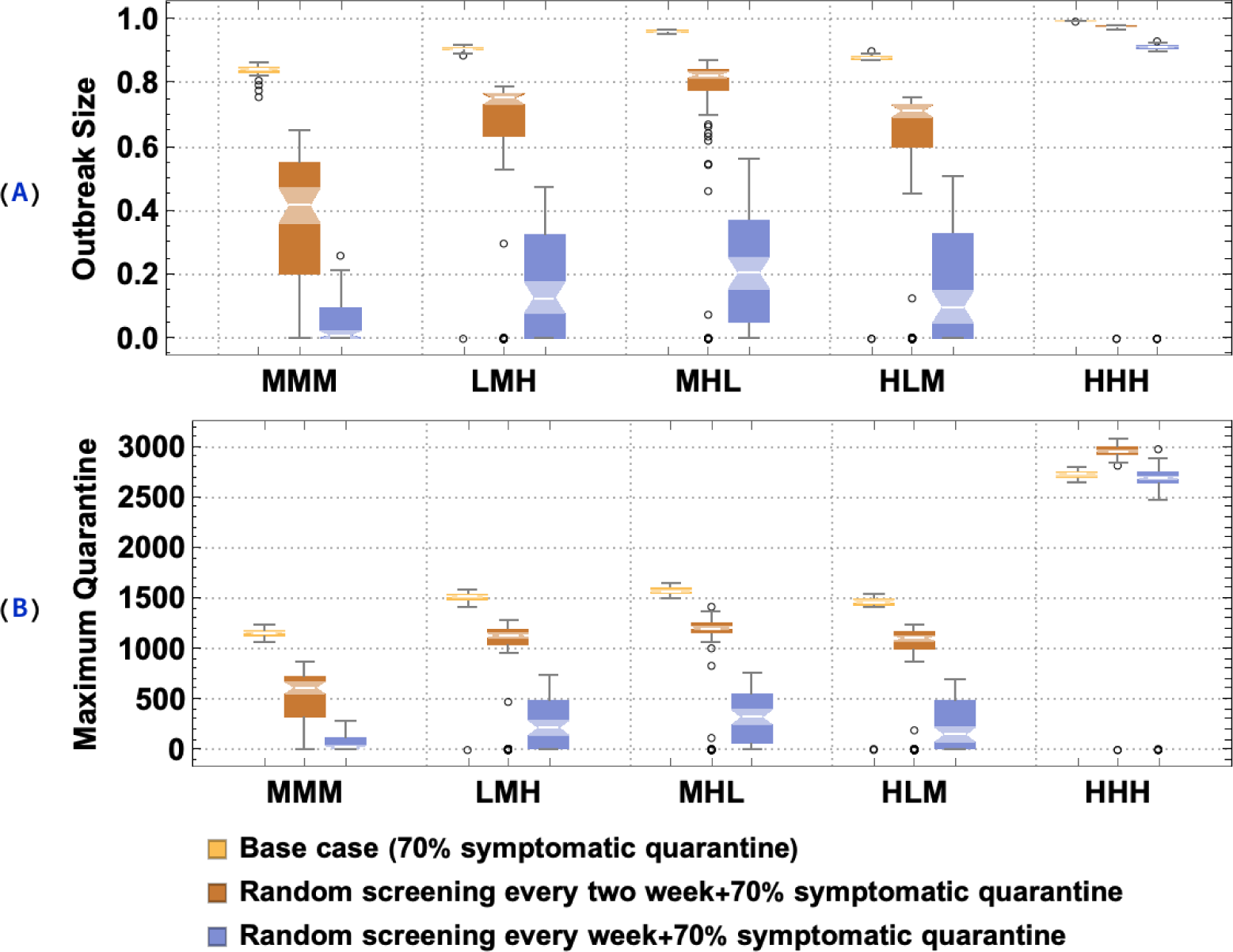
The effect of two different screening and test regulations on outbreak size and maximum quarantine capacity: random screening 100% of the population every two week and random screening 100% of population every week. We selected five typical cases as our baseline cases: MMM, LMH, MHL, HLM, and HHH, using the same setting as in Figure 7.

**Table 2:**
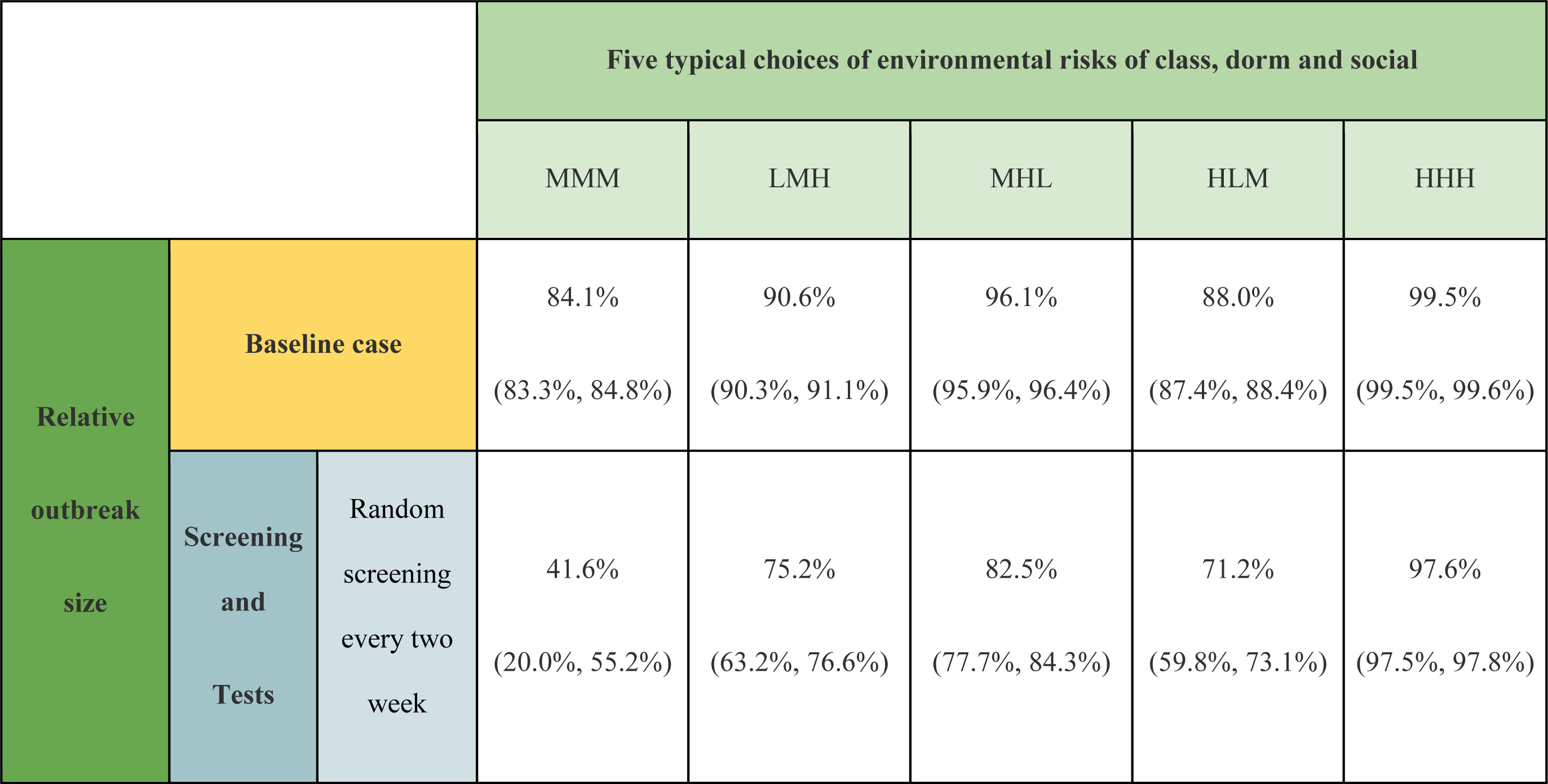
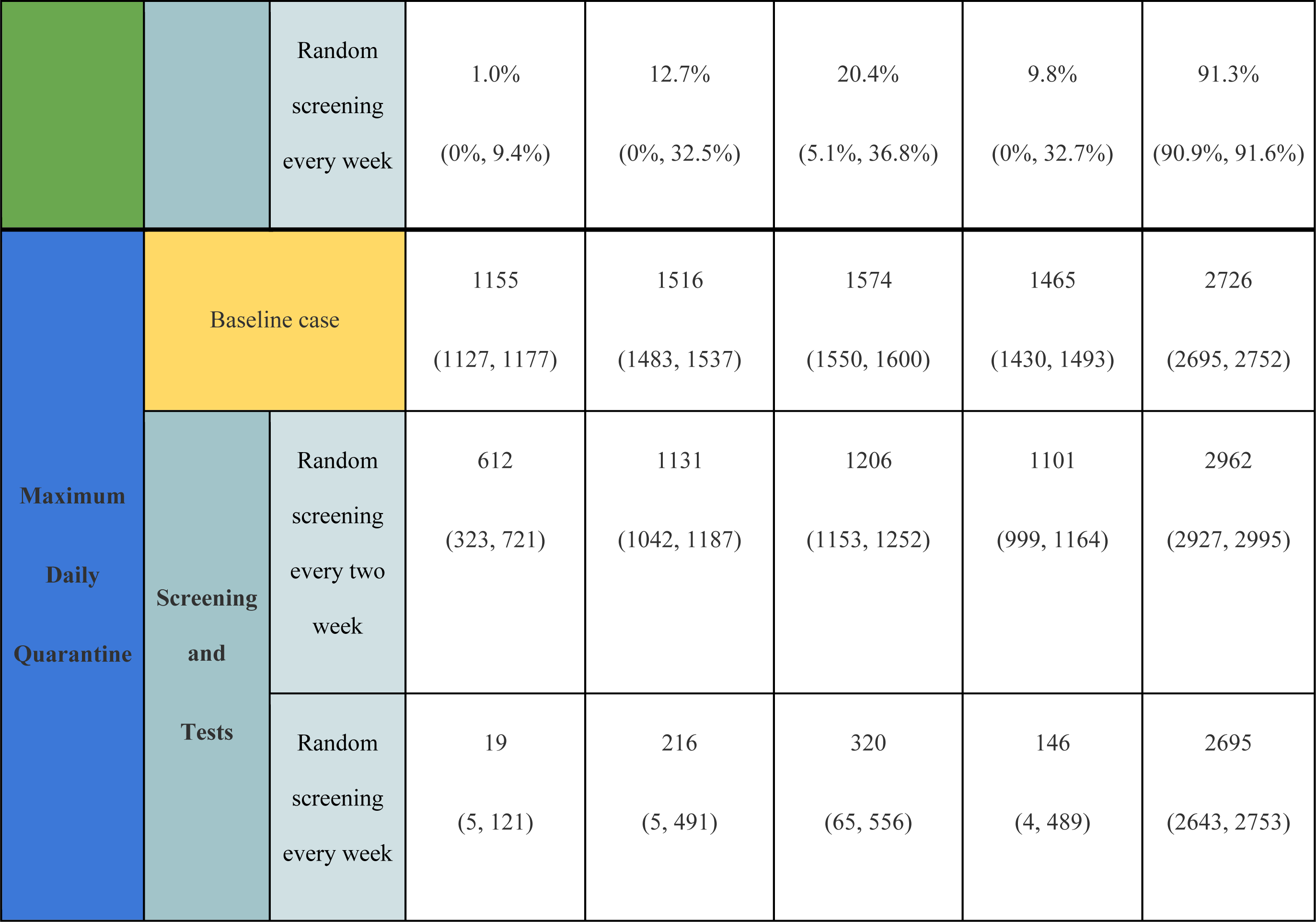
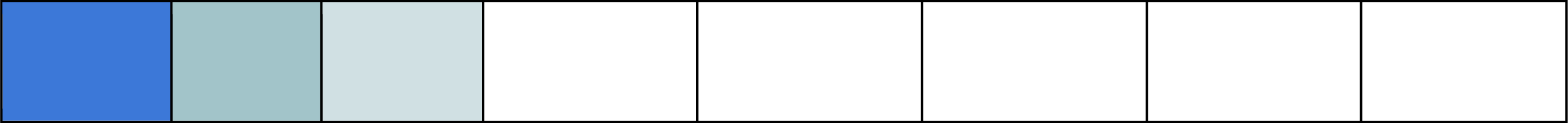
The effect of two different screening and test regulations on relative outbreak size and maximum quarantine capacity with a 6000 student body during a 16-week semester: random screening 100% of population every two week and random screening 100% of population every week. We selected five typical cases as our baseline cases: MMM (the first letter represents the risk level for class, followed by dorm and social), LMH, MHL, HLM, and HHH. The results shown are predicted median (25% quantile and 75% quantile).

#### Vaccination scenarios

Here we assume two simple vaccine scenarios (i) 50% students are vaccinated with 95% efficacy (ref), so we simply assume 50%*95%=47.5% students are fully protected (See Figure 10(A)); (ii) 80% students are vaccinated with 95% efficacy, i.e., 76% students are fully protected (See Figure 10(B)). We then explore the effect of lifting the screening and testing with less frequency, e.g., testing every week, every two weeks, every 4 weeks, every 8 weeks, and no random screening at all (i.e., symptomatic screening only). We selected five typical cases as our baseline cases: MMM (the first letter represents the risk level for class, followed by dorm and social), LMH, MHL, HLM, and HHH. A simple conclusion is that the herd immunity can be achieved as a result of 50% to 80% vaccination. When herd immunity is not reached, it is optimal to retain some level of environmental risk control on class, social and dorm (e.g., the use of facial masks). The impact of random screening was shown to be less important compared to environmental risk control.

**Figure 10:**
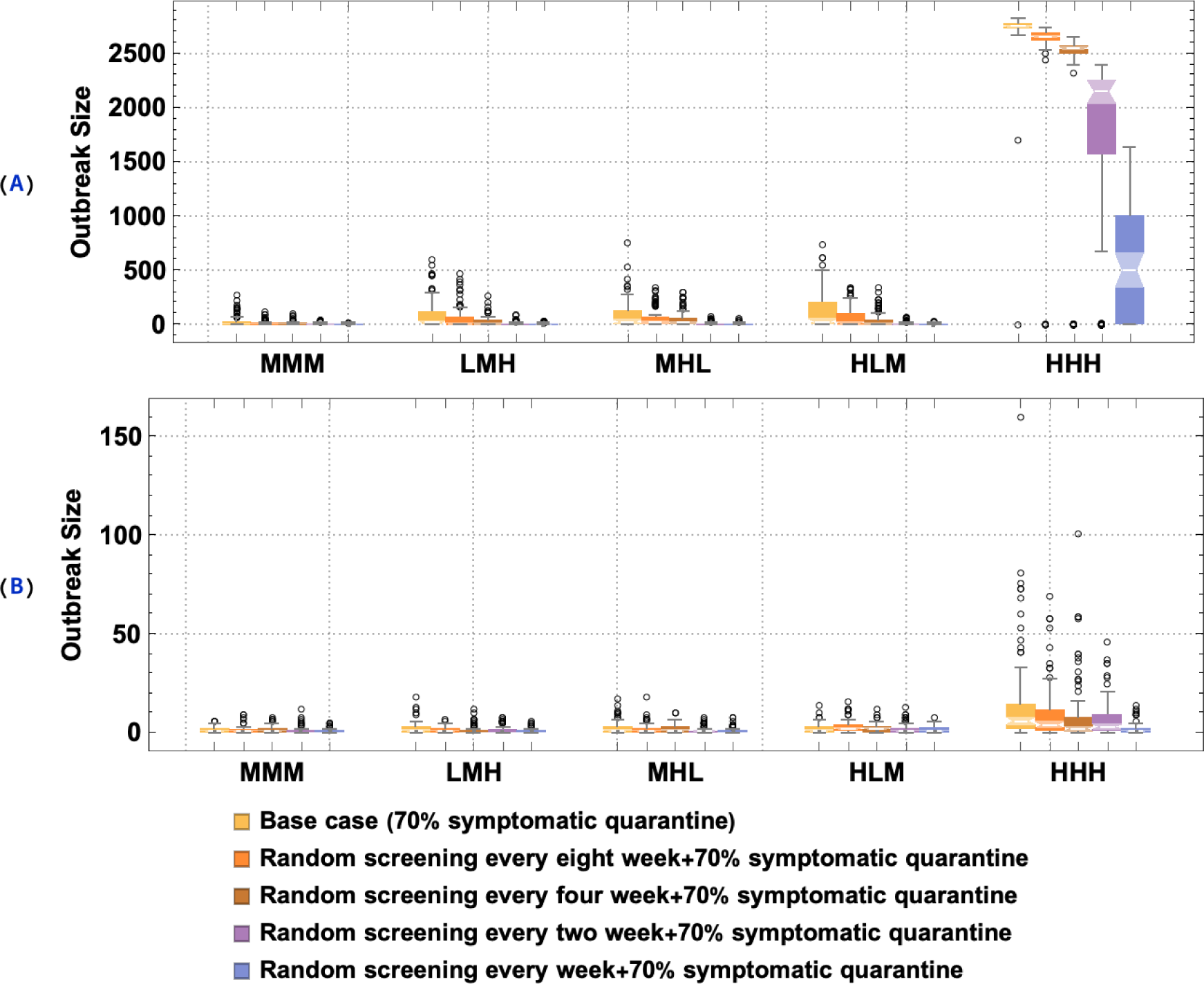
The effect of two vaccine scenarios and five different screening-and-test regulations on outbreak size and maximum quarantine capacity. Panel (A): 50% vaccinated with 95% efficacy; Panel (B): 80% vaccinated with 95% efficacy. For each vaccine scenario, we explore five different screening-and-test regulations: 1. no random screening (70% symptomatic quarantine); 2. random screening every eight week + 70% symptomatic quarantine; 3. random screening every four week + 70% symptomatic quarantine; 4. random screening every two week + 70% symptomatic quarantine; 5. random screening every week + 70% symptomatic quarantine; We selected five typical cases as our baseline cases: MMM, LMH, MHL,HLM, and HHH, using the same setting as in Figure 7.

## Discussion

Controlling the spread of COVID-19 is a critical challenge for colleges and universities planning to reopen on-campus educational, research, sports, and social activities in the fall. Even though the vaccination of college students has started on many college campuses across the US, it may not by itself prevent an outbreak due to incomplete coverage. University administrators are still wondering to what degree they should continue to implement non-pharmaceutical interventions and what are the most efficient control strategies.

To capture the disease dynamic on campus and answer those questions, we developed an individual-based model (IBM) of COVID-19 transmission that accounts for heterogeneity in social interactions, disease progression, and control interventions. Since social mixing plays an important part in COVID-19 transmission, we considered two basic types of mixing typical for college communities: (i) scheduled contacts (classes and dorm residence), (ii) random contacts (e.g., dining and/or other forms of unstructured social activity). Once infected each host undergoes a prescribed disease progression history, with a time-dependent infectivity pattern, viewed as a proxy for variable viral load during the course of infection. Such a computational framework allows us to generate and simulate more realistic college communities than standard SEIR models [8], and quantitatively assess the efficacy of different intervention strategies.

In our IBM setup, daily activities (class, dorm, and social mixing) takes place in a suitable environment, and each environment is assumed to have an associated risk factor (*a_c_*, *a_d_*, *a_s_*). These factors may mitigate the probability of transmission (exposure risk) in a given environment (0 < *a_i_* < 1). So they could represent site-specific control interventions (e.g., wearing facial masks, or providing clean contact space). Since these factors are highly uncertain and could vary in different colleges, we explore a range of choices of environmental risk factors, by taking three values (low, moderate, high) of each environmental risk factor (dorm, class, social mixing). Another control focus could be social activities (pool sizes). Our numeric simulations and analysis indicate that mixing-pool size could play an important role in containing COVID-19 outbreaks. We assessed the relative contribution of three environmental risk factors on outbreak size. Among these, the dorm risk was found as most significant, followed by classroom risk, and random social mixing. However, this observation is provisional on the underlying assumptions of the model about dorms, classrooms, social make-up.

A simple intervention based on symptomatic quarantine only was shown to have minimal impact on the outbreak size. The low efficacy of the symptomatic screening is due to the large contribution of asymptomatic hosts to disease transmission a delayed onset of symptoms, whereby pre-symptomatic (infective) individuals could still contribute to its spread. We found that the high frequency (weekly) random screening is the most effective non-pharmaceutical control strategy to mitigate and even prevent an outbreak, when coupled with an additional risk mitigation measure (e.g., facial mask) in classrooms, dorms, and social activities. Since random screening testing allows for the successful and early detection of both asymptomatic and symptomatic individuals, it successfully reduces the transmission rate such that the maximum quarantine capacity is far lower than expected to further reduce the economic burden caused from quarantine.

We also conducted a simple analysis on vaccination assuming 50% and 80% of the student body immunized with 95% vaccine efficacy. In most cases, it sufficed to prevent an outbreak or significantly mitigate its size. In highly transmissive environments, efficient environmental/behavioral control measures (i.e., wearing facial masks in class, dorms, or at social events) are required to mitigate disease spread even with a 80% vaccination coverage. But the frequency of random test screening could be highly reduced or replaced with symptomatic screening only (Figure 10).

Our results should be robust for colleges/universities with a smaller student body (See Figures S3-S8 in SI). The proposed individual-based modeling framework can be readily applicable to other aerosol or airborne infectious diseases like COVID-19. Indeed, the social mixing component of the system is independent of disease progress. The potential to generate and simulate synthetic college communities can be employed with robust predictions. Moreover, our methodology can be extended to other local community settings such as workplaces, primary and secondary schools, or hospitals.

Many additional topics could be explored by our IBM methodology, including consistent procedures to assess risk factors and relative contributions of different transmission environments and social mixing patterns on disease outbreak. Such assessment could guide effective intervention strategies. Our social mixing setup (scheduled and random) could be further expanded to account for more realistic intermediate levels of social connectivity. This work could also be extended to account for partial or dynamic vaccine schedules. Besides, since college campuses are often not isolated, we could extend current model to couple college campus transmission to the local community.

## Conclusions

Our analysis has implications for college reopening and in-person activities, planned in the near future. The risk of COVID-19 outbreak could be efficiently controlled or mitigated using existing control measures. Without vaccine, the high frequency (weekly) random screening appears the most effective control strategy, that would allow to mitigate and even prevent an outbreak on college campuses, coupled with some environmental/behavioral control (e.g., facial mask in class, dorm, and social events). Our analysis of vaccination shows that some level of herd immunity could be attained at 50% -80% vaccine coverage. Below herd immunity level, additional environmental/behavioral control measures are essential, but random screening is less important and could be reduced or lifted.

## Data Availability

N/A

## Acknowledgement

We would like to thank Dr. Daniel Tisch from the Department of Population and Quantitative Health Sciences, School of Medicine, Case Western Reserve University, for comments and discussions. We also would like to thank Audrey Tsoi, an undergraduate student from Department of Mathematics, Applied Mathematics and Statistics, Case Western Reserve University, for help with software and web implementation.

## Declarations of interest

None.

## Funding

This work was supported by the National Science Foundation RAPID Award [grant number DEB-2028631 to QH, AM, and DG] and the National Science Foundation RAPID Award [grant number DEB-2028632 to MNM]; Funders had no role in study design, writing of the report, or the decision to submit for publication. The corresponding authors had the final responsibility to submit for publication.

## Author contributions

**Qimin Huang:** Conceptualization, Methodology, Software, Writing - Original Draft, Visualization, Formal analysis; **Martial Ndeffo-Mbah:** Conceptualization, Methodology, Writing - Review & Editing; **Anirban Mondal:** Methodology, Writing - Review & Editing; **Sara Lee:** Resources, Writing - Review & Editing; **David Gurarie:** Conceptualization, Methodology, Software, Writing - Review & Editing, Supervision.

## Notes

### Competing Interest Statement

The authors have declared no competing interest.

