## Supplemental Information for "Individual-based modeling of COVID-19 transmission in college communities"

### Supplementary Information

#### Synthetic college community

The key inputs for a college community are student body size N, partitioned into two pools freshmen-sophomore, junior-senior with prescribed fraction ( $(f_f, f_s) = (0.4, 0.6)$. The partition determines their class-schedule, shared within each group, but no overlap between groups. Other contact activities (dorm, random social mixing) are shared equally among all students. Other key inputs include class-size distribution, weekly class schedules, dorm residence assignment, and random social mixing patterns. See table S2 for possible choices of parameters; but any other choices could be used to generate a synthetic college community.

##### Student body

We consider two types of colleges medium-size $(N = 6000)$, and small-size $(N = 3000)$.

**Classes schedule with seating** is generated by first calculating the number of classes offered $n_{class}$, according to the number of student $N$ and the mean class size $(ms_{class} = 50)$, i.e., $n_{class} \approx N / ms_{class}$. Classes are partitioned into MWF and TuTh schedules with a prescribe fraction of MWF, $w_{mwf}$. We also assign a time slot for each class on its scheduled days. In our setup, there were seven time slots available for MWF classes and five time slots available for TuTh classes. Every class is assigned a popularity (enrollment) weight distributed in the range [1, $w_{class}$], e.g., [1, 5]. Students are randomly assigned to three non-overlapping classes based on class-weights. The selection is done consistently to avoid class repetition and schedule conflict. In Figure S2, we show a typical distribution of class size, where we generate 137 classrooms for a 6000-student body.

**Seat Assignments in classroom:** We considered three sizes: 8 x 7 seats, 11 x 8 seats, and 13 x 10 seats. Each class was assigned to a sufficient classroom capacity greater than 1.2 times the class enrollment. Students were seated to optimize the distance between them.

##### Housing/Dorm

A randomly drawn fraction, $f_{dorm}$, of student body was assumed to reside in dorms, and socialize in dorm units. They were randomly partitioned into non-overlapping units of size $P_{dorm}$. Here we uses $P_{dorm} = 7$.

##### Random Social Mixing

Random social mixing pools are generated daily so that pool sizes $(m_1; m_2; m_3 ...)$ occur with frequencies $(f_1; f_2; ...)$ per host. The resulting mean contact rate per host is equal to $w = \sum_{m \geq 2} f_m m(m-1)$. In our simulations, we used two types of mixing patterns: (i) pool sizes (2, 3) with frequencies (⅔, ⅓). In this case, the mean contact rate (MCR) per host per day is equal to 3; (ii) pool sizes (2, 3, 4, 5) with frequencies (0.46, 0.31, 0.15, 0.08), where MCR=6. Such patterns could be viewed as a combination of personal social preferences and an imposed regulation of social mixing.

#### Infection and Disease Progress

Host infection upon contact (transition $S \rightarrow E$) is modeled as a Bernoulli random process with probability of success (infection) determined by the cumulative daily contacts of a given susceptible host. In each scheduled or random contact pool (class, dorm, social mixing), we estimate the survival probability (staying uninfected) depending on the pool's makeup, and associated-environmental risk factor. We assume each type of activity (class, dorm, and social) takes place in a suitable environment, and each environment has an associated mean *risk factor*

$(a_c, a_d, a_s)$. These factors $(0 < a_i < 1)$ mitigate the probability of transmission (exposure risk) in a given environment, They vary from 0 (full protection) to 1 (full exposure). In this form they could also be the target of control strategies. More generally, risk factors could be made site-specific (classroom and/or other place of activity). Hence, we explore a range of choices of environmental risk factors: three choices: low (0.1), moderate (0.2), and high (0.45) for three environmental risk factors (dorm, class, social risk);

Specifically, a contact pool with infectives $\{b_1;...;b_m\}$ (infectivity levels), and environmental risk factor $a$ gives survival probability $p_s = \prod_k (1 - ab_k)$. Hence, the survival probability from dorm and social mixing would be $p_{sd} = \prod_k (1 - a_d b_k); p_{ss} = \prod_k (1 - a_s b_k)$, respectively.

Classroom contacts differ from other environments, as the probability of survival depends on infectious status of nearest neighbors in adjacent seats, and the distance factor. Namely, $p_{sc} = \prod_k (1 - a_c d_k b_k)$, where distance factor $d_k$ decays as exponential function of distance $\vartheta_{ij}$ between seats $(i, j)$, $d_i = \exp(-\vartheta_{ij} / d)$ with scale factor $d$. In our simulation, we fixed $d = 2$.

The resulting probability of infection/ day is determined by cumulative daily contact pools of a given susceptible -host is given as $p = 1 - p_{sc} * p_{sd} * p_{ss,}$ where $p_{sc}, p_{sd}, p_{ss}$ are surviving probability from class, dorm and social contacts.

#### Model inputs

The college IBM was run on a daily basis for a 16-week semester. The key inputs to generate a synthetic college community in the model shown in Table S2 include: (i) student body inputs; (ii) class schedule inputs; (iii) dorm schedule inputs; (iv) social mixing inputs. The key inputs for disease progress in the model shown in Table S3 include: (i) fraction of the population that is in the symptomatic disease pathway; (ii) infectivity pattern for symptomatic and asymptomatic individuals; (iii) environmental risks for three environments (class, dorm, and social); (iv) pathogen spread length scale in classroom. All codes and simulations were implemented on Wolfram Mathematica platform.

#### Supplementary Tables

**Table S1: Contact pool size distribution.** mean contact rate/host $w = \sum_{m \geq 2} f_m m(m-1)$.

| | | | | |
|---|---|---|---|---|
| Contact pool size | 2 | 3 | 4 | … |
| Frequency/host | $f_2$ | $f_3$ | $f_4$ | … |

**Table S2. Inputs and parameters for a synthetic college community.**

| Generate a synthetic college community | | |
|---|---|---|
| **Parameter** | **Descriptions** | **Value** |
| **Student body** | | |
| $N$ | The total student body | 6000 (or 3000) |
| $(f_{fresh}, f_{senior})$ | Fraction of freshmen-sophomore, junior-senior students | (0.4,0.6) |
| **Class schedule inputs** | | |
| $ms_{class}$ | The mean class size | 50 |
| $w_{class}$ | The range of weight of class popularity | 5 |
| $w_{mwf}$ | The ratio of MWF classes to TuTh classes | 7/5 |
| **Dorm inputs** | | |
| $f_{dorm}$ | Fraction of student body that lives in dorms | 100% |
| $P_{dorm}$ | Maximum number of students allowed to share the hall, public utility, etc together | 7 |
| **Social mixing inputs** | | |
| $(m_1; m_2; m_3 ...)$ | Social mixing pool sizes | (2;3;4;5) |

| | | |
|---|---|---|
| $(f_1; f_2; f_3 ...)$ | The relative frequency of different pool sizes. | (0.46;0.31;0.15;0.08) |
| **Initial condition inputs** | | |
| $W$ | The number of weeks the model is run for | 16 |
| $I_0$ | The number of students who begin infected | 2 (or 1) |
| $f_{immune}$ | The fraction of the student body that begins immune | 0%, 50%, 80% |

**Table S3. Inputs and parameters for disease progress in college IBM.**

| **Disease progress** | | | |
|---|---|---|---|
| **Parameter** | **Descriptions** | **Value** | **Source** |
| $f_{symp}$ | Fraction of the population that is in the symptomatic disease pathway | 60% | [1] [2] |
| Infectivity pattern for Asymptomatic $\{b_{MA}, L_A, k_A\}$ | {Maximum infectivity of a person in the asymptomatic disease pathway, Duration in days of infection for a person in the asymptomatic disease pathway, Steepness} | {0.2, 12, 4} | [3] |
| Infectivity pattern for Symptomatic $\{b_{MS}, L_S, k_S\}$ | {Maximum infectivity of a person in the symptomatic disease pathway, Duration in days of infection for a person in the symptomatic disease pathway, Steepness} | {0.25, 21, 5} | [3] |
| $a_c$ | The environmental risk factor for class mixing | {0.1, .2, 0.45} low, moderate and high values | Assumption |
| $a_d$ | The environmental risk factor of dorm mixing | {0.1, .2, 0.45} | Assumption |

| | | | |
|---|---|---|---|
| | | low, moderate and high values | |
| $a_s$ | The environmental risk factor for social mixing | {0.1, .2, 0.45} low, moderate and high values | Assumption |
| $d$ | Pathogen spread length scale in classroom | 2 | Assumption |

#### Supplementary Figures

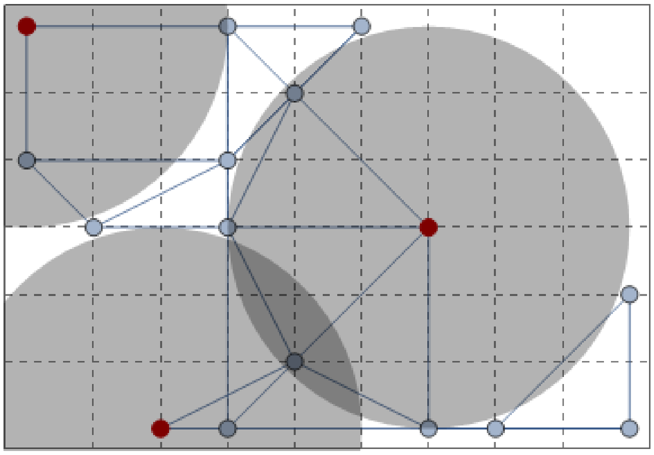

**Figure S1**: A typical sparsely seated classroom with 3 infected hosts (red) spreading pathogens within prescribed (shaded) areas. Graph links indicate nearest neighbors within distance < $r$, for all occupied seats (gray).

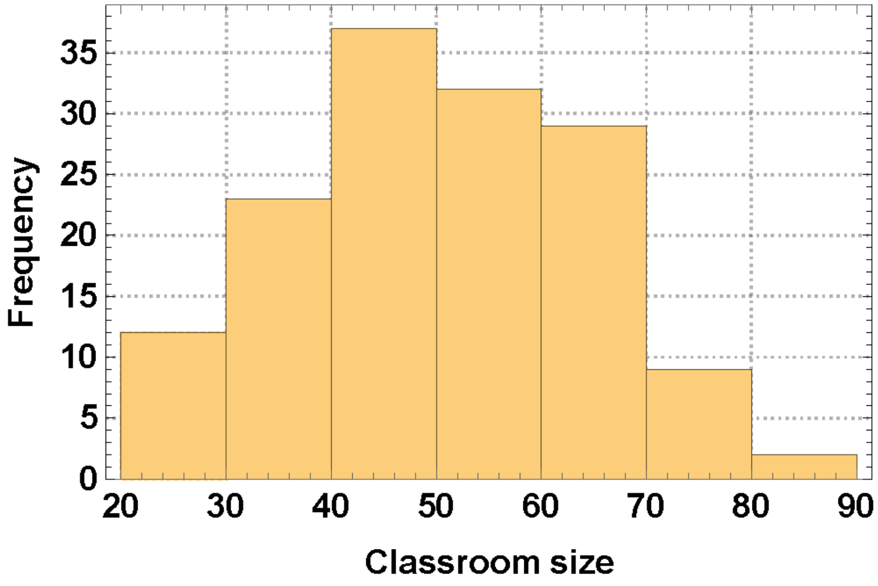


**Figure S2**: A typical distribution of class size for a 6000 student body, and 137 scheduled classes.

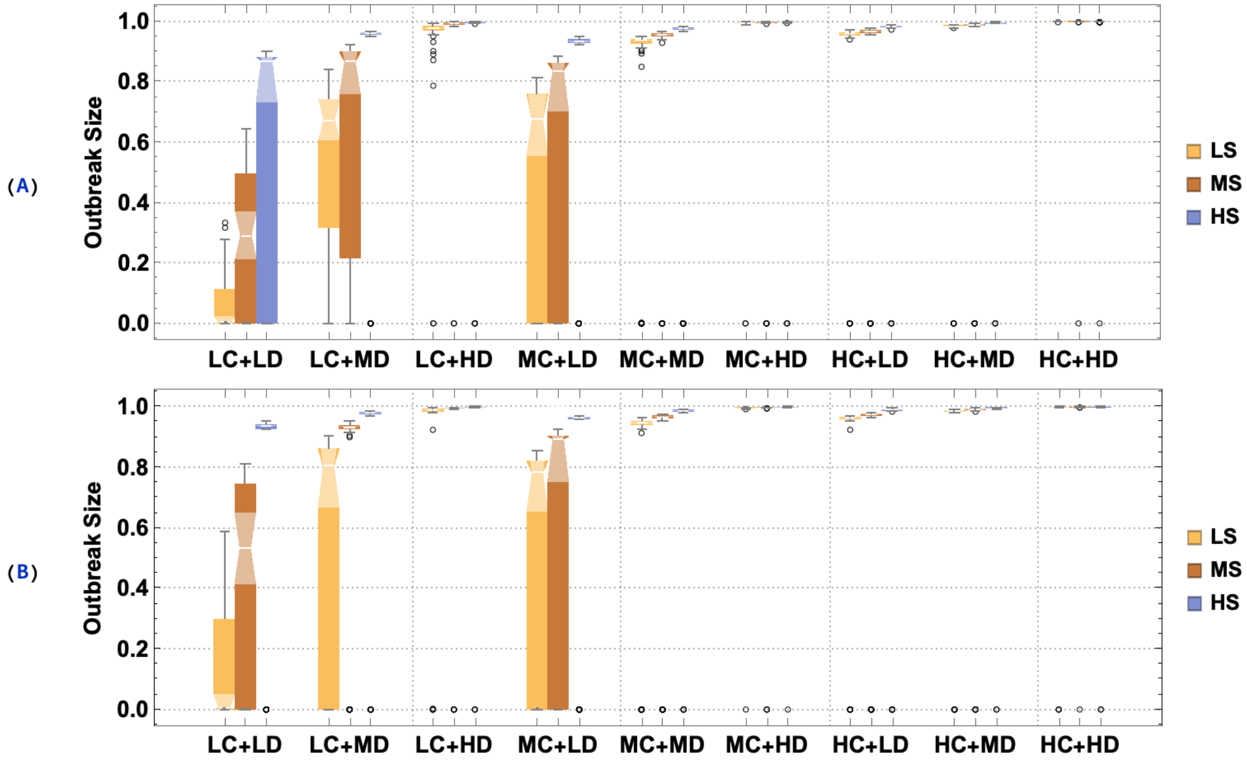


**Figure S3:** Relative outbreak-size distribution for 3 environmental risk factors (low, medium, and high) for three transmission environments (classroom, dorm, and random social mixing). The simulations were run over a 16-week (semester) period. Labels LC, MC, and HC represent low, medium, and high risk factors for class; LD, MD, and HD represent small, medium, and high risk factors for dorms; LS, MS, and HS represent small, medium, and high risk factors. The 27 parameter choices are split into triples. An ensemble of 100 realizations was run for each case. Panel (A) corresponds to a lower pool-size for random social mixing (mean contact rate MCR=3). Panel (B) corresponds to MCR=6 case. The student body size was $N = 3000$.

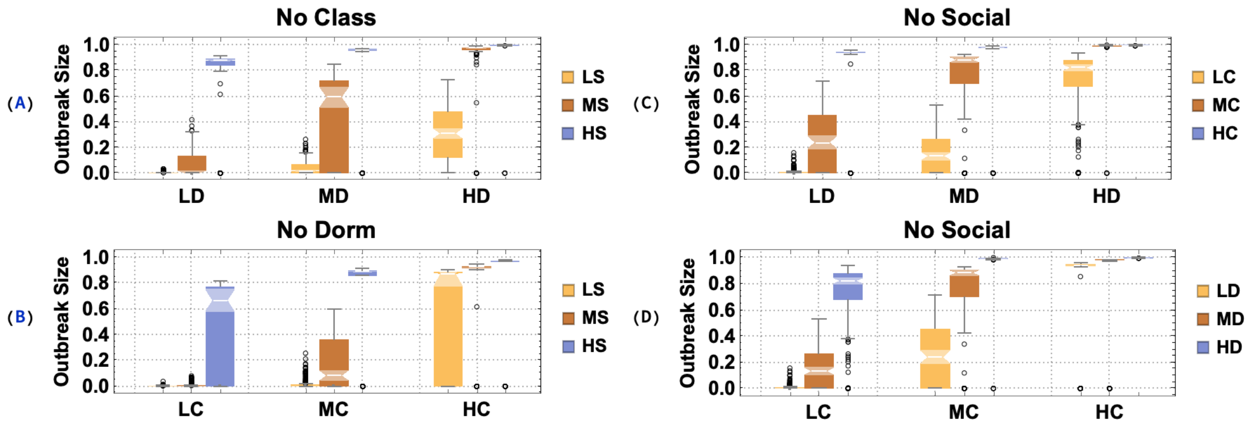


**Figure S4.** Same conditions and marking as Figure S3. Panel (A) shows the outbreak size distributions without class mixing (risk factor =0); Panel (B) shows the effect of reduced dorm mixing (risk factor =0). Panel (C) and (D) show the outbreak size distribution without random social mixing (risk factor =0). In all simulations MCR=6, and student body $N = 3000$.

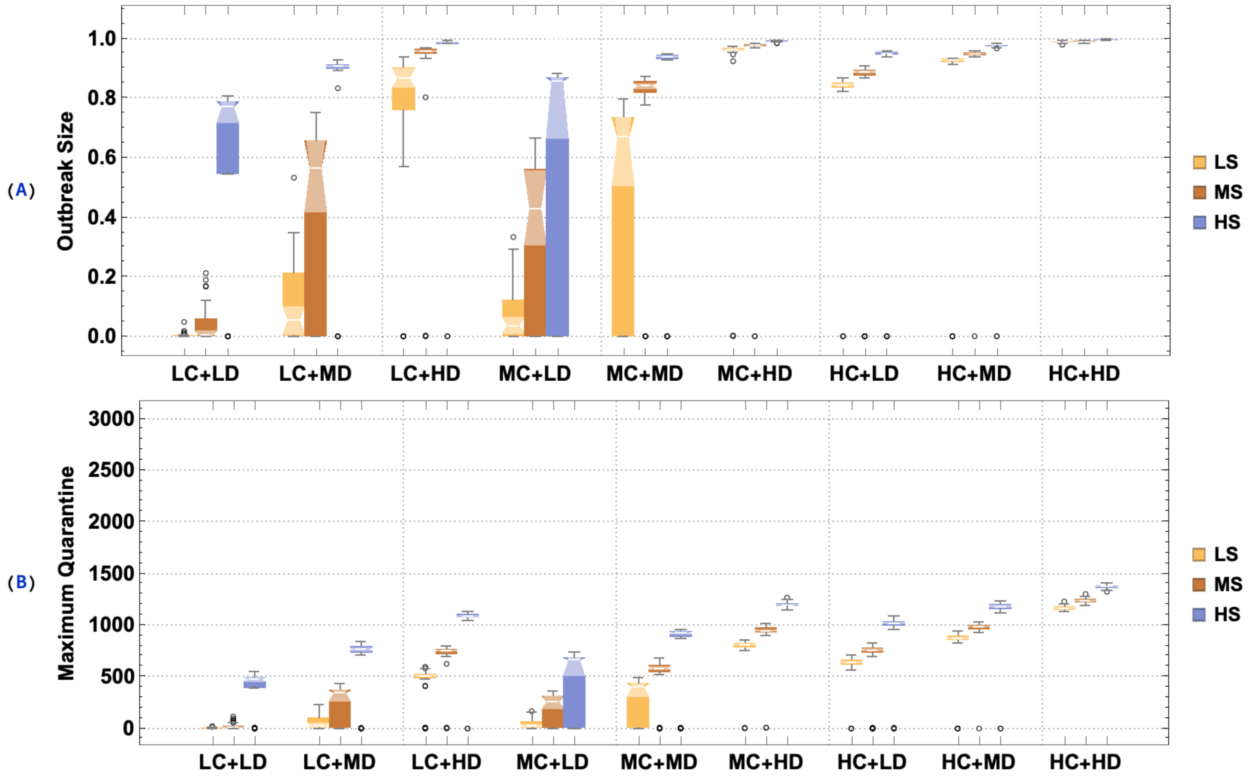


**Figure S5:** The effect of symptomatic quarantine on relative outbreak size and maximum quarantine capacity. An infected host exhibits symptoms around days 7, based on our time-dependent infectivity for the symptomatic pathway (Figure 2). 70% of randomly selected symptomatic hosts are put into quarantine.

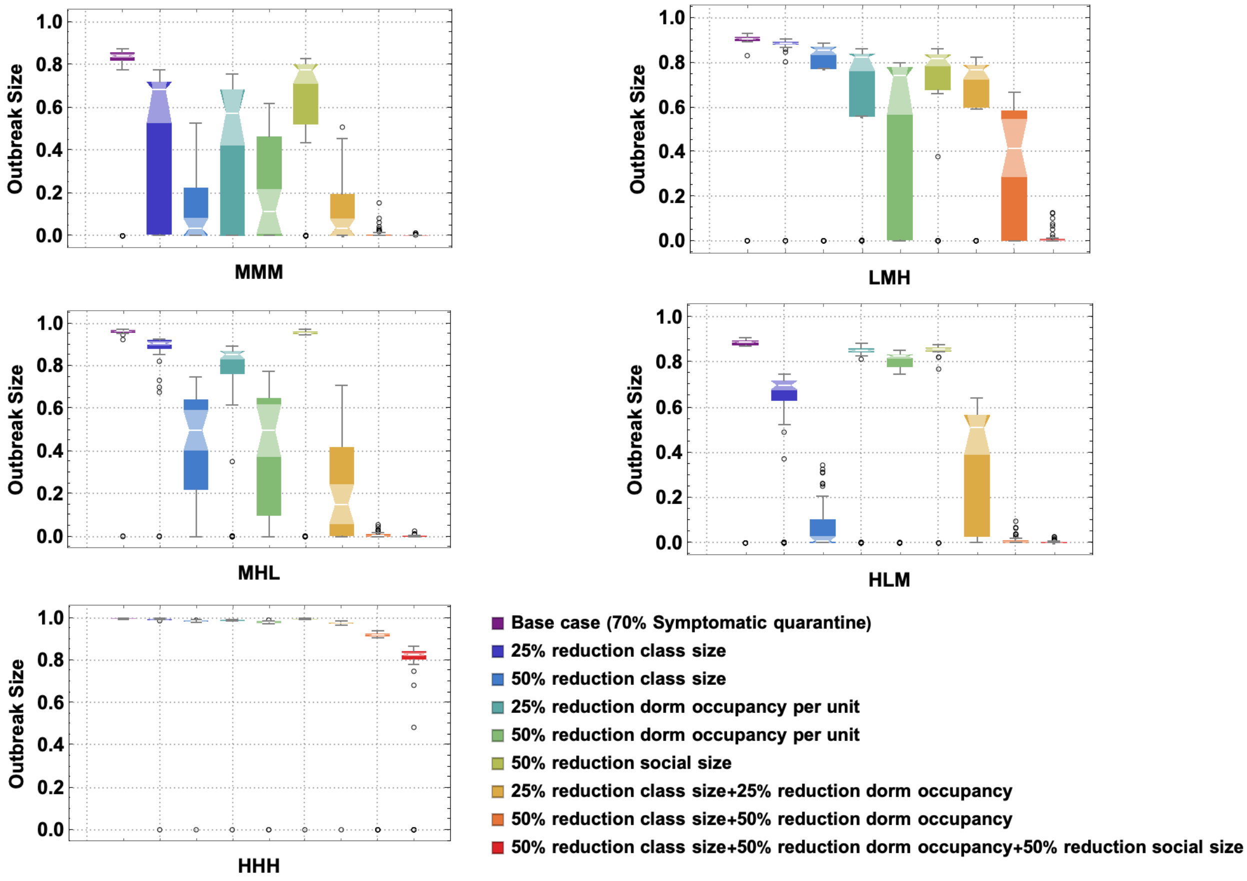


**Figure S6:** The effect of different combinations of class, dorm, and social mixing control on the outbreak size. Five baseline cases were selected for analysis with different choices of risk-factor (class, followed by dorm and social mixing), LMH, MHL, HLM, and HHH; LMH = low risk class, moderate dorm, and high risk social. We explored the 8 different control interventions of the (class, dorm, and social) size regulation.

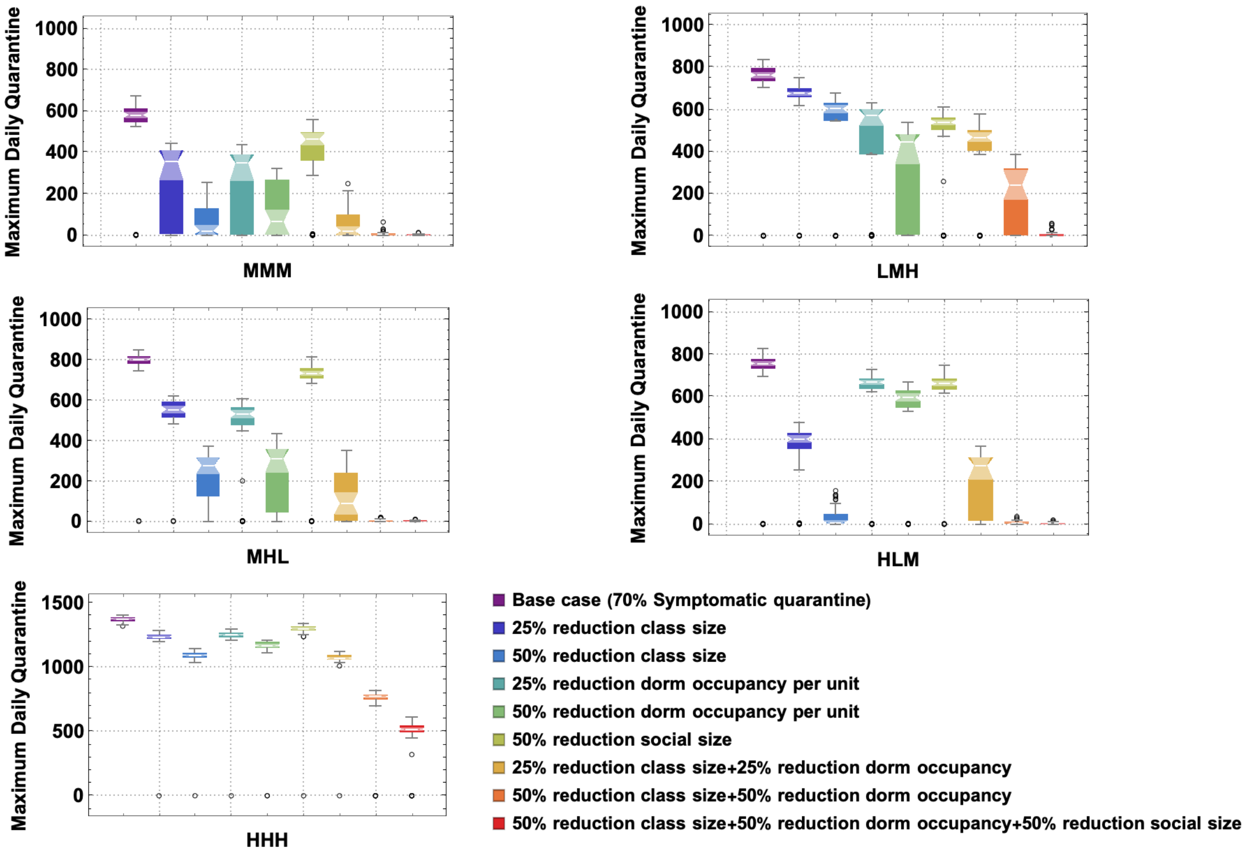


**Figure S7**: The effect of different combinations of class, dorm, and social control on maximum quarantine, using the same setting as Figure S6.

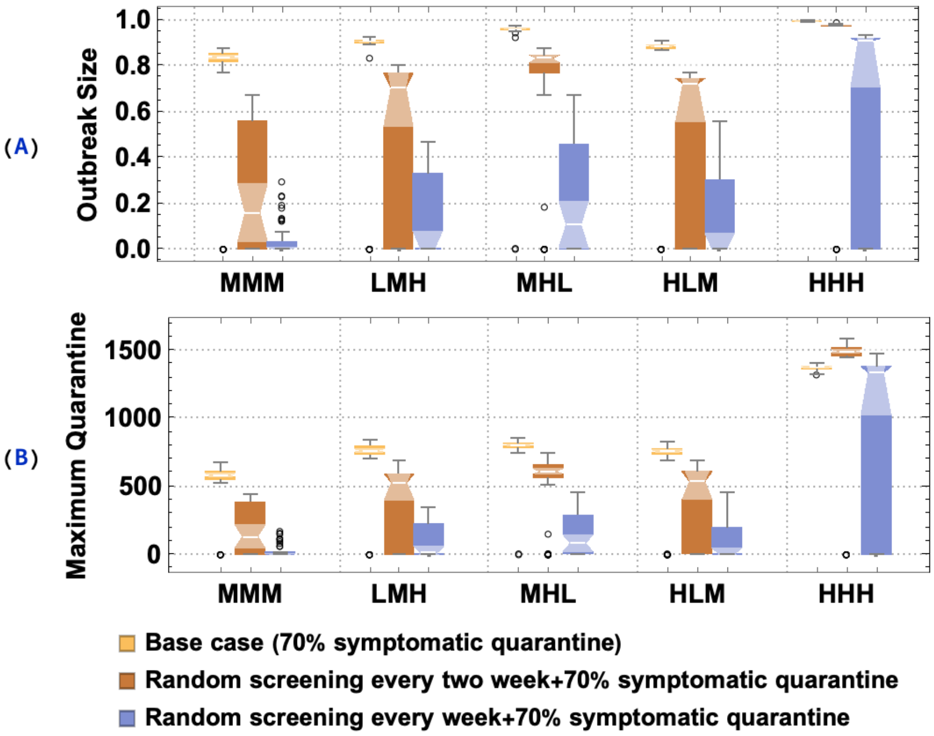


**Figure S8:** The effect of two test screening strategies on outbreak size and maximum quarantine capacity: random screening 100% of population every two week and randomly screening 100% of population every week. Five baseline cases were selected for analysis with different choices of risk-factor (class, followed by dorm and social mixing), LMH, MHL, HLM, and HHH; LMH = low risk class, moderate dorm, and high risk social.
